# A Scalable Biological Clock for Metabolic Disease Prediction from the Phenome India Cohort

**DOI:** 10.64898/2026.08.29.26361656

**Authors:** Pradeep Tiwari, Mayank Garg, Shrestha Pattanayak, Ishita Sarkar, Ritabrata Roy, Naveen Bhatraju, Anshul Verma, Sreeshma Raj K, Satyartha Prakash, Vignesh S Kumar, Mohammad Azhar Uddin, Nancy Rawat, Ankita Sahu, Yogesh Kumar, Pulkit Hasmukhbhai Leuva, Ankita Mridha, Vamsi Yenamandra, Ajay Pratap Singh, Aastha Mishra, Swasti Raychaudhuri, Karthik Bharadwaj Tallapaka, Giriraj Ratan Chandak, Mahesh J Kulkarni, Mahesh Dharne, Romi Wahengbam, Jatin Kalita, Prasenjit Manna, Umakanta Subudhi, Saikat Majumder, Partha Chakraborty, Kumardeep Chaudhary, Shantanu Sengupta, Phenome India Consortium, Viren Sardana, Shilpak Chatterjee, Dipyaman Ganguly

**Affiliations:** Department of Biology, Trivedi School of Biosciences, Ashoka University, Sonipat, Haryana 131029, India; Koita Centre for Digital Health, Trivedi School of Biosciences, Ashoka University, Sonipat, Haryana 131029, India; Datla Human Immunome Lab, Trivedi School of Biosciences, Ashoka University, Sonipat, Haryana 131029, India; Division of Cancer Biology and Inflammatory Disorders, CSIR-Indian Institute of Chemical Biology, Kolkata, West Bengal, India; CSIR-Institute of Genomics and Integrative Biology, New Delhi 110007, India; Academy of Scientific and Innovative Research, Ghaziabad 201002, India; CSIR-Centre for Cellular and Molecular Biology, Hyderabad, Telangana 500007, India; Biochemical Sciences Division, CSIR-National Chemical Laboratory, Pune, Maharashtra 411008, India; Center for Infectious Diseases, Biological Sciences and Technology Division, CSIR-North East Institute of Science and Technology, Jorhat, Assam 785006, India; CSIR-Institute of Minerals and Materials Technology, Bhubaneswar, Odisha 751013, India; Division of Infectious Diseases and Immunology, CSIR-Indian Institute of Chemical Biology, Kolkata, West Bengal 700032, India; Division of Cell Biology and Physiology, CSIR-Indian Institute of Chemical Biology, Kolkata, West Bengal 700032, India

## Abstract

**Background:** India has a rising incidence of chronic non-communicable diseases, making it a major healthcare burden today. Growing evidence suggests that chronic low-grade inflammation links ageing with cardiometabolic disorders, captured by the emerging concept of inflammaging. However, most evidence on biological ageing comes from Western populations, with no similar models developed for the Indian population. Given the country’s distinctive genetic makeup, unique exposome, and heterogeneous NCD presentation, Western models may not capture inflammaging and its effects in the Indian population.

**Methods:** We analysed baseline data from 4,240 adults in the Phenome India CSIR Health Cohort Knowledgebase (PI CheCK), a nationwide multi-centre cohort. Participants were stratified into eight cardiometabolic phenotype groups by BMI (Asian cut off), blood pressure and HbA1c status. We trained a Super Learner ensemble to predict chronological age in the lean normotensive-normoglycaemic reference group (n=615) using 44 plasma cytokines, sex, haemoglobin, and bioimpedance-derived visceral fat area, per cent body fat, and total body water. Performance was assessed by repeated five-fold cross-validation and in a held-out healthy test set. Calibrated biological age acceleration was then estimated in the remaining 3,625 participants.

**Results:** Median age was 51.0 years (IQR 41.0 to 62.0) and 49.4% were female. The Super Learner outperformed elastic net and XGBoost comparators. Permutation importance identified visceral fat area, per cent body fat, CTACK, SDF1a, haemoglobin and sex as leading contributors, with body composition measures accounting for the largest share, indicating an immune-metabolic rather than cytokine-only signal. Biological age acceleration was concentrated in overweight/obese phenotypes. Lean phenotypes showed acceleration close to the reference (0.32 0.50 years).

**Conclusions:** Cytokine and body composition measures capture a quantifiable immunometabolic ageing signal in a South Asian cohort, with acceleration driven predominantly by adiposity. External validation and longitudinal follow up are required.

## Introduction

Non-communicable diseases (NCDs) now account for 75% of all deaths worldwide, and cardiometabolic disorders including type 2 diabetes, prediabetes and hypertension contribute greatly to this burden [1]. NCDs contribute disproportionately to premature mortality in low-and middle-income countries [1]. For example, in India, cardiovascular disease onset occurs roughly a decade earlier than in Western populations, and the country now harbors the world’s second-largest number of adults with diabetes (an estimated 89.8 million in 2024, projected to approach 157 million by 2050 [2–4]. Nationally representative surveys also provide a very similar picture, e.g., the ICMR-INDIAB study reported that a substantial fraction of Indian adults already have diabetes or prediabetes and that roughly one in four have hypertension, with district-and state-level analyses confirming wide, and still poorly explained, geographic heterogeneity in hypertension prevalence and control across the country [5, 6]. This burden is further compounded by a distinctive South Asian phenotype of central, visceral adiposity at comparatively low body-mass index, which confers cardiometabolic risk that risk models derived from European or American cohorts often do not capture well [7, 8].

Chronic, sterile, low-grade inflammation, designated as ‘inflammaging’, has emerged as a plausible mechanism that links aging to this cluster of age-related diseases [9–11]. Inflammaging is not a fixed, universal process but rather a lifelong, individually sculpted trajectory that begins in utero and is shaped by the cumulative interplay of genetics, infections, diet, adiposity, gut-barrier integrity, cellular senescence and socioeconomic circumstances [9]. Concurrent anti-inflammatory (‘anti-inflammaging’) mechanisms often buffer its pathophysiological consequences, from tissue damage and disease risk to adaptive homeostasis. However, these vary markedly between individuals and, importantly, between populations and socioeconomic strata [9, 12].

In recent years, studies have examined different inflammaging-specific biological clocks across populations worldwide, though more frequently in the Western world. For instance, the ‘iAge’ model, derived from 50 cytokines, chemokines and growth factors using a guided autoencoder, tracks multimorbidity, immunosenescence, frailty and cardiovascular aging [9, 13]. Other efforts, such as ‘ipAge’, refined this approach with 38 markers in a linear model, and ‘SImAge’ used a deep-learning algorithm to reduce the panel to ten analytes[9, 14, 15]. Proteomic and epigenetic–inflammatory hybrid models such as ‘ProtAge’ and ‘EpInflammAge’ extend this effort further, linking inflammation-adjacent markers to incident chronic disease and all-cause mortality[9, 16, 17].

These clocks have two major limitations to their usefulness for population-scale screening, particularly in the LMIC countries. First, despite convergent performance, the specific marker sets of these different clock models overlap only minimally, and most constituent analytes require costly, technically demanding assays [9]. Second, and most consequentially for the present study, virtually all of these clocks have been trained and validated in European, North American, or Han Chinese/Russian cohorts. The very few studies that have looked beyond this, such as in the Yakutian population of extreme-cold Siberia, revealed that inflammaging signatures and their age-accelerated components are not universal but shift with ethnicity, climate, and environment [9, 18, 19].

The Indian population, constituting one-sixth of humanity, is genetically and environmentally distinct from the cohorts underlying most existing biological clocks. To our knowledge, India has no reported biological clock customized for its population. The most comprehensive recent review in the field explicitly calls for ‘a standardized set of inflammaging markers for first-line screening’ that could be deployed routinely in large aging populations, complemented as needed by more specific and expensive assessments [9]. This is precisely the gap the present study addresses in the Indian context.

The Phenome India-CSIR Health Cohort Knowledgebase (PI-CheCK) provides a suitable setting to develop population-relevant biological age models for Indian adults because it combines standardized cardiometabolic phenotyping, body-composition assessment, hematological and biochemical measurements, and plasma cytokine profiling in a nationwide cohort[20, 21].

In this study, we leverage baseline data from the PI-CheCK cohort to derive a low-cost, scalable biological clock model of inflammaging with a set of markers chosen through successive iterations, specifically to optimize for low per-sample cost, its reliance on instrumentation and assays already deployable at scale in resource-constrained settings, and its accessibility outside specialized research laboratories. Our model was derived from plasma cytokine abundance data, a limited set of routine clinically relevant hematological and biochemical analytes, as well as body-composition indices captured by multi-frequency bioimpedance analysis, which offers a similarly low-cost, field-deployable means of assessing central adiposity, which is mechanistically and epidemiologically central to cardiometabolic risk in South Asians[7, 8]. We also evaluated whether the final minimal model for measuring age acceleration is associated with, and predictive of, prediabetes, type 2 diabetes, and hypertension among Indian adults. In doing so, we aim to provide a scalable, population-appropriate tool for identifying individuals at accelerated cardiometabolic-inflammatory risk within India’s rapidly evolving NCD landscape, adaptable to other resource-limited settings that currently lack representation in the biological-clock literature.

## Methods

### Phenome-India Cohort

The Phenome India-CSIR Health Cohort Knowledgebase (PI-CheCK) is a nationwide, prospective, multi-center cohort of employees, retirees, and spouses of the Council of Scientific and Industrial Research. The cohort recruited 10,267 participants across 37 laboratories and associated centers spanning 27 cities, 17 states, and two union territories. Baseline data collection was completed between December 2023 and June 2024. Participants underwent standardized anthropometry, body-composition assessment, and hematological and biochemical testing. Plasma cytokine profiling was performed in a subset of participants[20].

### Study Population And Cardiometabolic Phenotype Classification

The working cohort included participants with available circulating cytokine measurements, anthropometric data, blood pressure phenotype data, and glycemic measurements. Participants were classified according to three cardiometabolic axes: BMI status, blood pressure status, and glycemic status.

BMI status was defined using the Asian BMI cutoff, with participants classified as lean if BMI was <25 kg/m2 and overweight/obese if BMI was >=25 kg/m2. Hypertension was defined as systolic/diastolic blood pressure >=140/90 mmHg, self-reported history of high blood pressure, or use of antihypertensive medication; participants not meeting these criteria were classified as normotensive. The glycemic status was defined using HbA1c, with HbA1c >=5.7% classified as hyperglycemic and HbA1c <5.7% classified as normoglycemic.

We combined the three binary classifications to generate eight cardiometabolic phenotype groups. The lean normotensive-normoglycemic group was used as the healthy reference cohort. We randomly divided this reference cohort into a healthy training set and an independent healthy test set. All remaining phenotype groups were used as the cardiometabolic validation cohort.

### Data Preprocessing

Cytokines with detectable non-zero values in more than 30% of the healthy reference cohort were retained for downstream analysis. Four cytokines with low detection frequency were excluded, resulting in 44 cytokines for modeling. Exact zero cytokine values were interpreted as values below the assay detection or reporting limit and were replaced with a small constant value of 0.001 prior to model fitting.

### Exploratory Cytokine Analyses

We performed descriptive analyses to characterize cytokine variation across age, sex, and cardiometabolic phenotype groups. For each retained cytokine, Spearman correlation was used to evaluate the association between cytokine concentration and chronological age. False discovery rate correction was applied across cytokines. To assess whether cytokine-age associations were consistent by sex, Spearman correlations were estimated separately in females and males and compared graphically. In addition, phenotype-associated cytokine differences were evaluated using linear models with log2-transformed cytokine concentration as the outcome and cardiometabolic phenotype group as the primary predictor, adjusting for age and sex. The lean normotensive-normoglycemic group was used as the reference group.

### Model Development and Cross-Validation Framework

Chronological age was modeled in the healthy reference cohort using 44 cytokines, sex, total body water, visceral fat area, percent body fat, and hemoglobin as predictors. Waist-hip ratio was excluded to reduce redundancy with adiposity-related phenotype definitions.

Model performance was evaluated using repeated cross-validation within the healthy reference cohort. Five-fold cross-validation was repeated five times. In each repeat, the healthy reference cohort was divided into five folds; models were trained on four folds and evaluated on the held-out fold. This procedure generated out-of-fold predicted biological age estimates for healthy reference participants. Model performance was summarized using R², mean absolute error, and root mean squared error.

### Super Learner Biological Age Model

A Super Learner ensemble was used as the primary biological age prediction model. Super Learner combines predictions from multiple candidate algorithms and estimates the optimal weighted combination of these learners using cross-validation [22, 23]. The learner library included ordinary least-squares regression, ridge regression, LASSO, elastic-net regression, random forest, ranger random forest, XGBoost, and a mean-model benchmark.

For each cross-validation training split, each candidate learner was trained and used to generate internal cross-validated predictions. Ensemble weights were then estimated within the training portion of each split by minimizing mean squared prediction error. We constrained weights to be non-negative and to sum to one. The held-out fold was not used during model fitting or ensemble-weight estimation. The final Super Learner prediction was calculated as the weighted average of predictions from the candidate learners.

XGBoost was implemented conservatively to reduce overfitting, using shallow trees, shrinkage, subsampling, column subsampling, and regularization. The number of boosting rounds was selected using internal cross-validation within the training data.

### Model Evaluation

Model performance was evaluated using repeated 5-fold cross-validation within the healthy reference cohort. In each repeat, the cohort was partitioned into five folds; models were trained on four folds and evaluated on the held-out fold. This procedure was repeated across five independent repetitions. Performance was summarized using R², mean absolute error, and root mean squared error. XGBoost [24] and elastic-net regression [25] were evaluated as individual comparator models using the same repeated cross-validation framework.

### Final Model Fitting and Application

After cross-validation-based model evaluation, we trained a final Super Learner model using the healthy training set. This final model was evaluated in the independent healthy test set and was subsequently used for biological age prediction in the cardiometabolic validation cohort. Predictor-contribution analysis was performed for the final fitted model using model-agnostic permutation importance in the independent healthy test set. For each predictor, values were randomly permuted and the resulting increase in prediction error was used to quantify its contribution to model performance.

### Biological Age Calibration and Age Acceleration

To account for systematic bias and age-related compression in predicted biological age estimates, predicted biological age was calibrated against chronological age within the healthy reference cohort [26, 27]. Calibration was performed using out-of-fold predicted biological age values generated during repeated cross-validation, ensuring that calibration was based on predictions from models that had not been trained on the corresponding individuals.

Expected predicted biological age was estimated as a function of chronological age in the healthy reference cohort. Biological age acceleration was then calculated as the difference between predicted biological age and expected predicted biological age. Positive values indicated higher predicted biological age than expected for chronological age, whereas negative values indicated lower predicted biological age than expected.

After model development, the final Super Learner model was trained using the healthy reference cohort and applied to the cardiometabolic validation cohort. The calibration equation derived from the healthy reference cohort was then applied to the validation cohort to calculate calibrated biological age acceleration. Group-wise mean biological age acceleration was summarized across cardiometabolic phenotype groups.

### Variable Importance Analysis

Variable importance for the Super Learner model was evaluated using a model-agnostic permutation approach. Because the Super Learner combines predictions from multiple candidate algorithms, variable importance was assessed by quantifying the reduction in predictive performance after perturbing each predictor rather than by relying on model-specific coefficients [28].

Permutation importance was estimated within the repeated cross-validation framework. For each held-out fold, baseline prediction performance was first calculated using the unmodified held-out data. Each predictor was then randomly permuted within the held-out fold, while all other predictors were left unchanged, and predicted biological age was recalculated. Variable importance was defined as the increase in prediction error after permutation compared with the baseline prediction error. Importance values were averaged across folds and repeats.

Predictors with larger increases in prediction error after permutation were interpreted as contributing more strongly to biological age prediction. Because correlated predictors may share predictive information, permutation importance was interpreted as predictive contribution to the ensemble model rather than evidence of causal biological relevance [29].

### Application to Cardiometabolic Phenotype Groups

After model development in the healthy reference cohort, the final Super Learner biological age model was applied to participants in the cardiometabolic validation cohort. This cohort included all participants outside the lean normotensive-normoglycemic reference group. Predicted biological age was generated for each validation participant using the same predictor set as in the healthy reference model.

The calibration equation derived from the healthy reference cohort was then applied to the validation cohort to estimate expected predicted biological age for each participant’s chronological age. Biological age acceleration was calculated as the difference between observed predicted biological age and expected predicted biological age. Mean calibrated biological age acceleration was then summarized across the seven cardiometabolic phenotype groups and compared with the healthy reference group.

This analysis was used to determine whether cardiometabolic phenotypes showed evidence of higher biological age than expected for chronological age relative to the healthy reference pattern.

## Results

### Cohort Characteristics and Phenotype Stratification

The analytic cohort included 4,240 participants. Baseline characteristics across cardiometabolic phenotype groups are summarized in **Table 1**. The overall median age was 51.0 years (IQR, 41.0-62.0), and 2,095 participants (49.4%) were female. Median BMI was 26.3 kg/m² (IQR, 24.0-29.1), and median HbA1c was 5.7% (IQR, 5.3-6.3). Participants in overweight/obese phenotype groups had higher visceral fat area and percent body fat than lean phenotype groups. Based on the Asian BMI cutoff, 1,455 participants were classified as lean (BMI <25 kg/m²) and 2,785 as overweight/obese (BMI ≥25 kg/m²). Hypertension was identified in 1,919 participants, while 2,321 were normotensive. Hyperglycemia was present in 2,141 participants and 2,099 were normoglycemic.

**Table 1.** Baseline characteristics of the analytic cohort by cardiometabolic phenotype group. Continuous variables are presented as median (interquartile range), and categorical variables are presented as n (%). Cardiometabolic phenotype groups were defined using BMI status, blood pressure status, and glycemic status. The healthy reference group comprised lean normotensive-normoglycemic participants. Abbreviations: OW/OB, overweight/obese; HTN, hypertensive; NT, normotensive; HG, hyperglycemic; NG, normoglycemic; IQR, interquartile range; BMI, body mass index; HbA1c, glycated hemoglobin; HDL, high-density lipoprotein; LDL, low-density lipoprotein.

| Characteristic | Overall | OW/OB<br>HTN-HG | OW/OB<br>HTN-NG | OW/OB<br>NT-HG | OW/OB<br>NT-NG | Lean<br>HTN-HG | Lean<br>HTN-NG | Lean<br>NT-HG | Healthy<br>Ref |
| --- | --- | --- | --- | --- | --- | --- | --- | --- | --- |
| N | 4240 | 890 | 483 | 607 | 805 | 350 | 196 | 294 | 615 |
| Age, years | 51.0 (41.0-62.0) | 59.0 (52.0-67.0) | 52.0 (45.0-61.0) | 51.0 (43.0-59.0) | 42.0 (36.0-48.0) | 65.0 (57.0-72.0) | 57.0 (48.0-67.0) | 54.5 (46.0-63.8) | 40.0 (34.0-46.5) |
| Female sex, n (%) | 2095 (49.4) | 456 (51.2) | 251 (52.0) | 325 (53.5) | 480 (59.6) | 114 (32.6) | 80 (40.8) | 112 (38.1) | 277 (45.0) |
| Current smoking, n (%) | 213 (5.0) | 40 (4.5) | 26 (5.4) | 32 (5.3) | 33 (4.1) | 23 (6.6) | 10 (5.1) | 23 (7.8) | 26 (4.2) |
| BMI, kg/m<sup>2</sup> | 26.3 (24.0-29.1) | 28.7 (26.8-31.5) | 28.3 (26.4-30.6) | 28.1 (26.4-30.5) | 27.4 (26.0-29.6) | 23.4 (22.0-24.1) | 23.1 (21.9-24.1) | 23.0 (21.6-24.1) | 22.9 (21.5-24.0) |
| HbA1c, % | 5.7 (5.3-6.3) | 6.4 (6.0-7.3) | 5.4 (5.2-5.5) | 6.1 (5.8-6.8) | 5.3 (5.1-5.5) | 6.3 (5.9-7.3) | 5.4 (5.2-5.5) | 6.1 (5.8-6.9) | 5.2 (5.0-5.4) |
| Total cholesterol | 176.0 (150.5-202.9) | 171.2 (143.8-201.6) | 180.2 (154.8-206.8) | 179.0 (152.2-206.3) | 179.7 (155.8-202.3) | 170.1 (140.8-204.2) | 180.2 (156.5-211.4) | 176.2 (151.0-205.8) | 174.3 (151.2-195.2) |
| HDL cholesterol | 48.0 (41.6-54.7) | 47.6 (41.3-54.5) | 48.5 (41.8-55.2) | 47.1 (41.1-53.8) | 47.5 (41.2-53.3) | 49.2 (42.9-56.1) | 51.8 (43.7-59.7) | 49.1 (42.0-56.2) | 47.8 (42.8-55.5) |
| LDL/HDL cholesterol ratio | 2.40 (1.94-2.89) | 2.35 (1.86-2.87) | 2.45 (2.03-2.93) | 2.51 (2.02-3.00) | 2.51 (2.13-2.94) | 2.22 (1.72-2.75) | 2.33 (1.89-2.81) | 2.40 (1.93-2.88) | 2.32 (1.90-2.80) |
| Total body water | 31.3 (27.0-36.4) | 31.7 (27.2-37.7) | 32.4 (27.8-37.2) | 31.5 (27.3-37.2) | 31.2 (27.3-37.3) | 30.8 (25.8-34.1) | 30.8 (25.6-34.2) | 30.4 (25.3-34.8) | 30.5 (26.2-35.3) |
| Visceral fat area | 172.0 (123.2-229.0) | 236.5 (188.0-300.8) | 208.0 (166.0-265.0) | 207.0 (165.0-253.0) | 167.0 (134.0-214.0) | 151.0 (118.5-182.0) | 126.0 (101.0-165.0) | 122.0 (99.0-158.8) | 99.0 (76.0-129.0) |
| Percent body fat, % | 36.6 (30.7-43.1) | 41.3 (35.5-46.2) | 40.4 (33.4-45.5) | 40.3 (34.1-45.3) | 39.6 (33.2-44.2) | 31.0 (27.1-36.0) | 30.8 (26.5-36.1) | 30.5 (26.1-36.1) | 29.8 (26.0-35.1) |
| Waist-hip ratio | 0.99 (0.93-1.06) | 1.06 (1.01-1.12) | 1.03 (0.98-1.08) | 1.03 (0.98-1.08) | 0.98 (0.94-1.03) | 0.98 (0.93-1.02) | 0.94 (0.90-1.00) | 0.94 (0.90-0.98) | 0.89 (0.84-0.94) |
| Hemoglobin | 13.2 (12.1-14.3) | 13.0 (12.0-14.0) | 13.2 (12.2-14.4) | 13.1 (12.0-14.3) | 13.0 (11.9-14.3) | 13.3 (12.3-14.4) | 13.2 (12.2-14.6) | 13.4 (12.2-14.3) | 13.4 (12.3-14.7) |

Continuous variables are shown as median (IQR); categorical variables are shown as n (%). OW/OB = overweight/obese; HTN = hypertensive; NT = normotensive; HG = hyperglycemic; NG = normoglycemic; Healthy Ref = lean normotensive-normoglycemic reference group.

Combining BMI, blood pressure, and glycemic status generated eight cardiometabolic phenotype groups. The lean normotensive-normoglycemic group served as the healthy reference cohort and included 615 individuals. The remaining 3,625 participants comprised the cardiometabolic validation cohort. Among participants with overweight/obesity, 890 were hypertensive-hyperglycemic, 483 were hypertensive-normoglycemic, 607 were normotensive-hyperglycemic, and 805 were normotensive-normoglycemic. Among lean participants, 350 were hypertensive-hyperglycemic, 196 were hypertensive-normoglycemic, and 294 were normotensive-hyperglycemic. **Figure 1A-F** shows the distribution of age, sex, BMI, percent body fat, and hemoglobin across phenotype groups.

**Figure 1.**
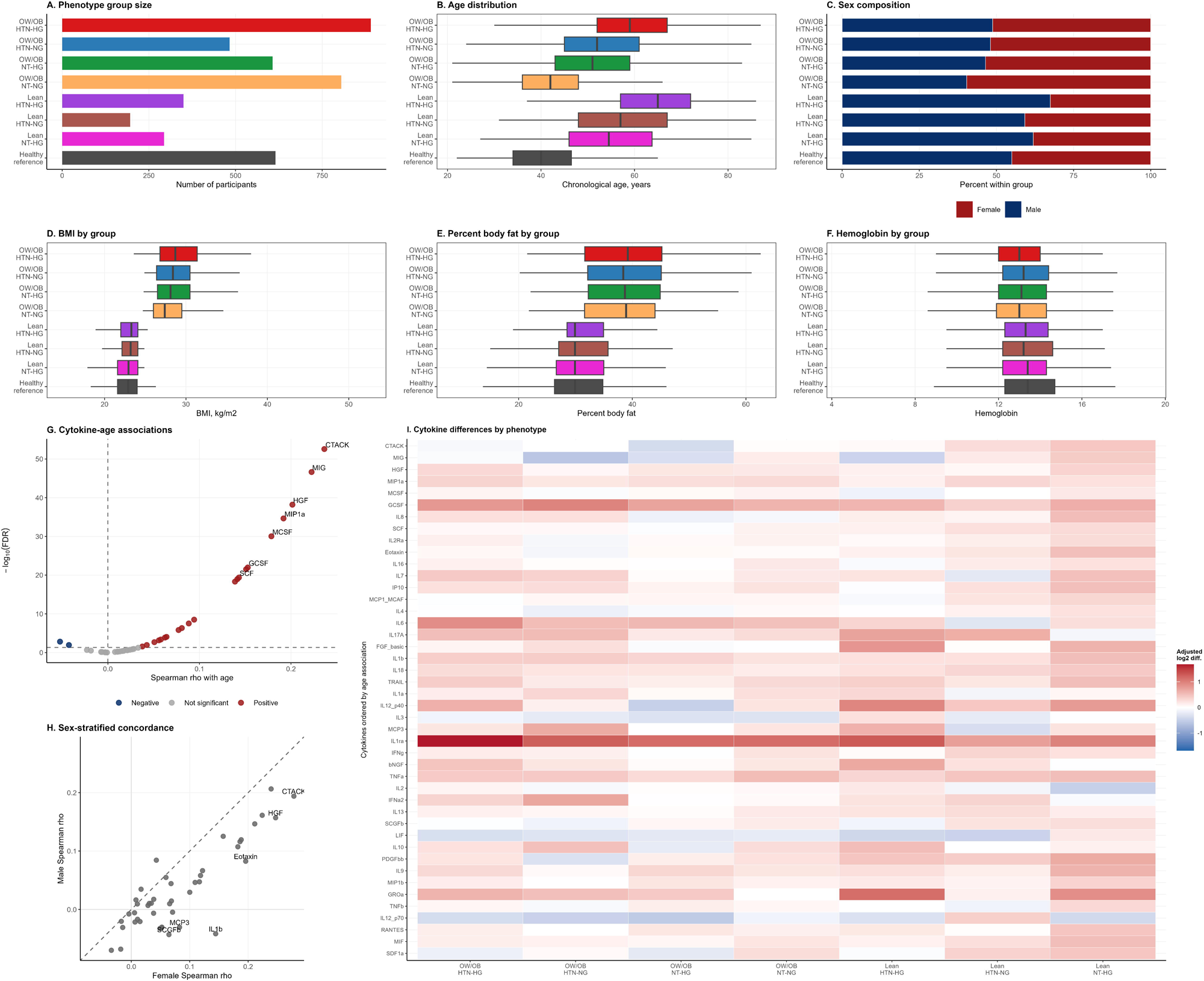
Cohort structure and exploratory inflammaging landscape. (A) Distribution of participants across the eight cardiometabolic phenotype groups defined by BMI status, blood pressure status, and glycemic status. (B) Chronological age distribution across phenotype groups. (C) Sex composition within each phenotype group. (D-F) Distribution of BMI, percent body fat, and hemoglobin across phenotype groups. (G) Volcano plot showing Spearman correlations between chronological age and each of the 44 retained cytokines, with false discovery rate correction across cytokines. (H) Sex-stratified concordance of cytokine-age correlations; each point represents one cytokine, with female-specific Spearman rho on the x-axis and male-specific Spearman rho on the y-axis. The dashed diagonal indicates equal age association in females and males. (I) Heatmap showing age-and sex-adjusted cytokine differences across cardiometabolic phenotype groups relative to the lean normotensive-normoglycemic healthy reference group. Cytokines are ordered by their overall age association. Abbreviations: OW/OB, overweight/obese; HTN, hypertensive; NT, normotensive; HG, hyperglycemic; NG, normoglycemic.

Cytokine-age associations were evaluated across all 44 retained cytokines. Several cytokines showed modest but significant positive correlations with chronological age, with CTACK, MIG, HGF, MIP1a, and MCSF showing the strongest associations (**Figure 1G; Supplementary Table S3**). Cytokine-specific scatter plots are provided in **Supplementary Figure S2**. Sex-stratified analyses showed broad concordance of cytokine-age correlations between females and males, although some cytokines showed sex-specific variation in effect size (**Figure 1H; Supplementary Table S4**). Age-and sex-adjusted comparisons across cardiometabolic phenotype groups showed broad cytokine differences relative to the healthy reference cohort, with prominent differences in IL1ra and IL6 across metabolically adverse groups (**Figure 1I; Supplementary Table S5**).

### Model Development And Cross-Validated Performance

We evaluated the performance of the inflammation-associated biological age model using repeated 5-fold cross-validation within the healthy reference cohort. The Super Learner model showed the highest overall predictive performance, achieving a mean cross-validated R² of 0.356, mean absolute error of 6.83 years, and root mean squared error of 8.93 years (**Figure 2A-C; Supplementary Table S1**). In comparison, elastic-net regression achieved a mean R² of 0.326, mean absolute error of 6.95 years, and root mean squared error of 9.12 years, while XGBoost achieved a mean R² of 0.299, mean absolute error of 7.21 years, and root mean squared error of 9.32 years (**Supplementary Table S1**).

**Figure 2.**
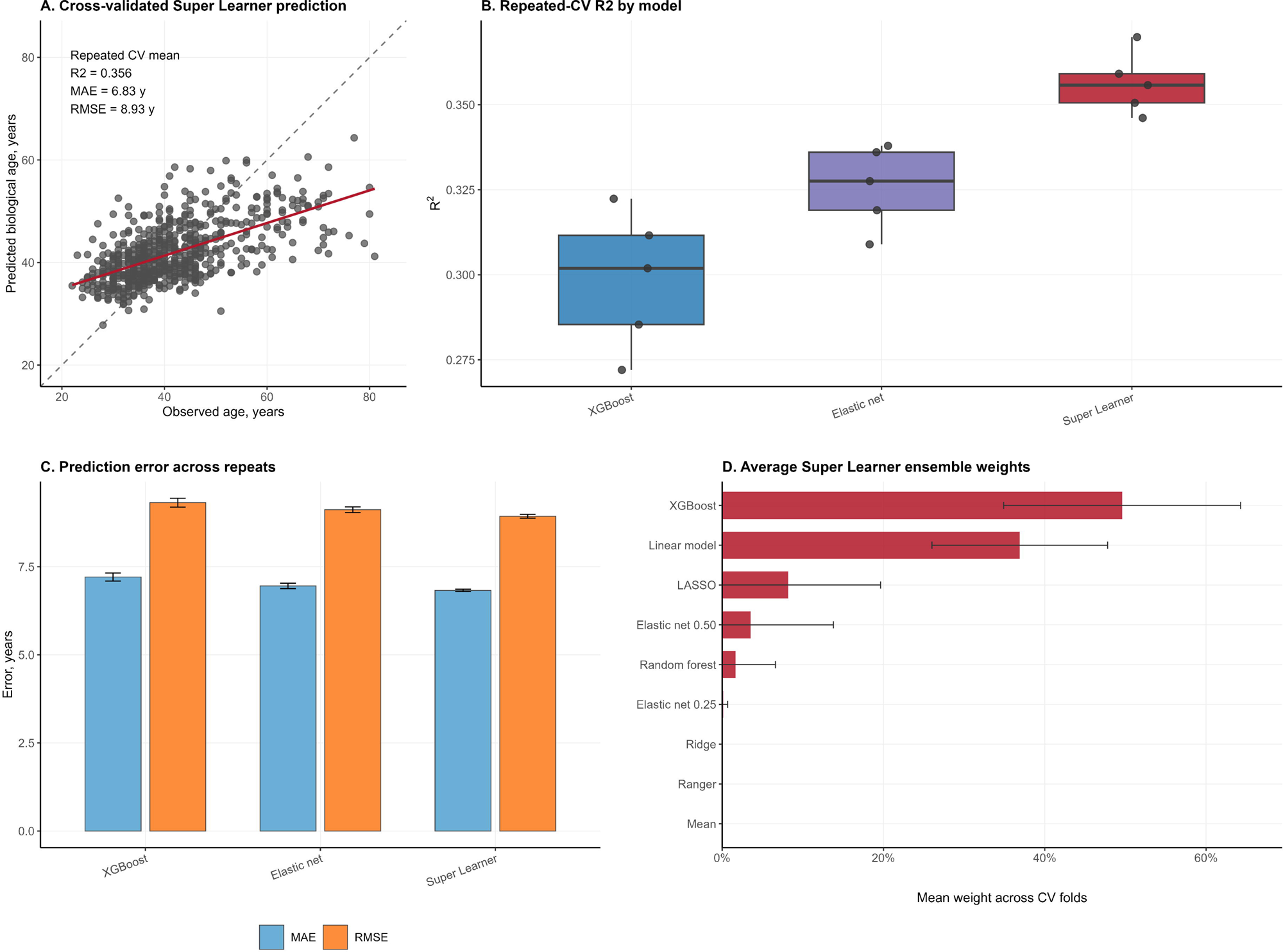
Development and cross-validation performance of the Super Learner biological age model. (A) Cross-validated Super Learner predictions of biological age versus observed chronological age in the healthy reference cohort. Points represent participant-level predictions averaged across repeated cross-validation runs. (B) Repeated 5-fold cross-validation R² across XGBoost, elastic-net regression, and Super Learner models. (C) Mean absolute error and root mean squared error across repeated cross-validation runs. (D) Average Super Learner ensemble weights across cross-validation folds. Abbreviations: CV, cross-validation; MAE, mean absolute error; RMSE, root mean squared error.

The Super Learner ensemble assigned the largest average weight to XGBoost, followed by ordinary least-squares regression and LASSO regression (**Figure 2D; Supplementary Table S2**). Mean ensemble weights were 0.496 for XGBoost, 0.369 for ordinary least-squares regression, and 0.082 for LASSO, with smaller contributions from elastic-net and random forest models. These findings indicate that the ensemble combined both nonlinear and linear predictive components and achieved modestly better age prediction than the individual comparator models.

After cross-validation, a final Super Learner model was trained using the healthy training set and evaluated in the independent healthy test set. In this held-out test set, the final model achieved an R² of 0.501, mean absolute error of 6.24 years, and root mean squared error of 8.20 years. This final fitted model was used for predictor-contribution analysis and for application to the cardiometabolic validation cohort.

### Predictor Contributions to the Biological Age Model

Predictor contribution was assessed using model-agnostic permutation importance in the held-out healthy test set. The final Super Learner model achieved an R² of 0.501, MAE of 6.24 years, and RMSE of 8.20 years. Ensemble weights were concentrated on XGBoost and ordinary least-squares regression, indicating that both nonlinear and linear components contributed to prediction. Permutation analysis identified visceral fat area, percent body fat, CTACK, SDF1a, hemoglobin, and sex as the strongest contributors to model performance (**Figure 3A; Supplementary Table S6**). When summarized by predictor class, body-composition variables accounted for the largest share of positive permutation importance, followed by cytokines **(Figure 3B**). These findings suggest that the model captured an immune-metabolic aging signal rather than a cytokine-only inflammatory signal.

**Figure 3.**
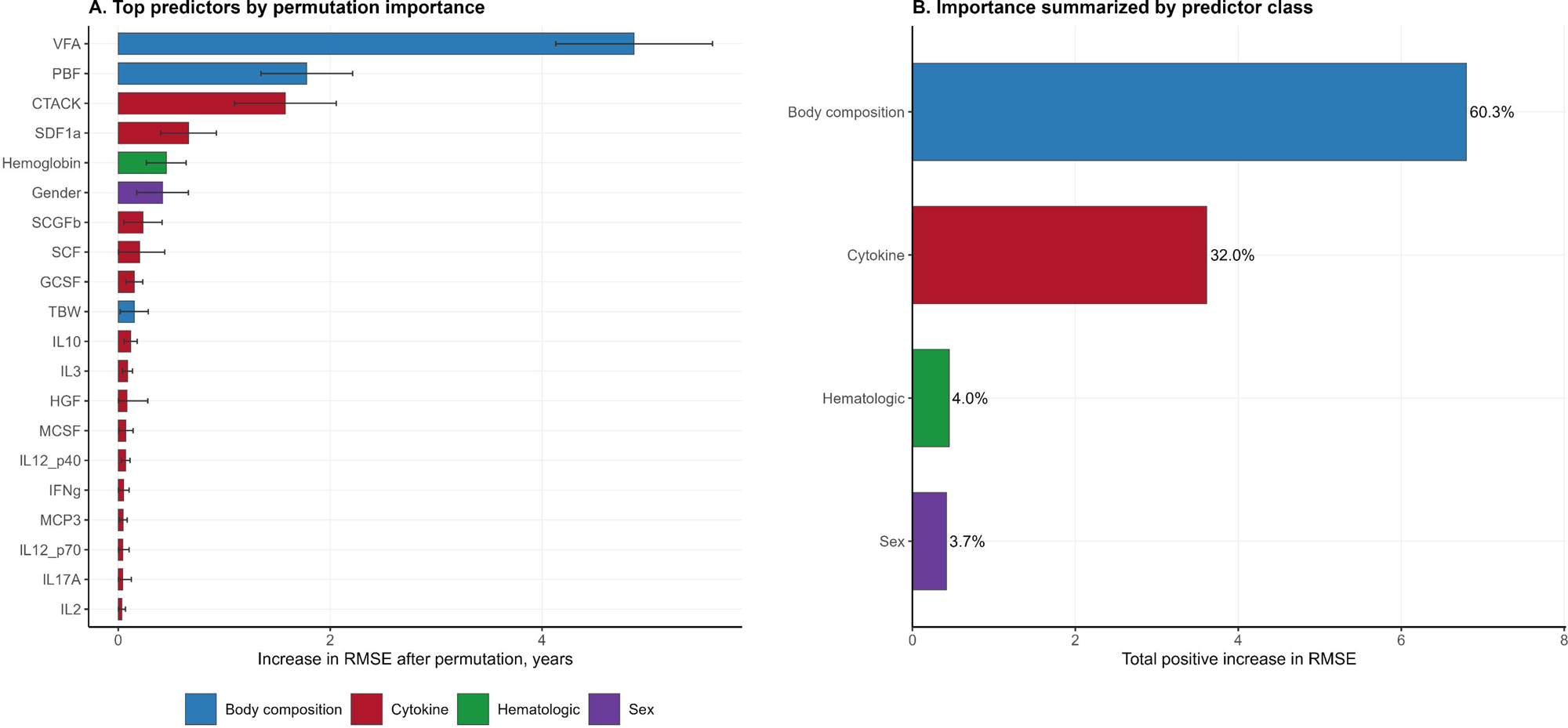
Predictor contributions to the Super Learner biological age model. (A) Top predictors ranked by model-agnostic permutation importance in the final fitted Super Learner model. Importance is shown as the increase in root mean squared error after permuting each predictor, with larger values indicating greater contribution to biological age prediction. Error bars represent variability across permutation iterations. (B) Predictor importance summarized by feature class, calculated as the total positive increase in root mean squared error across predictors within each class. Percentages indicate the relative share of total positive permutation importance. Abbreviations: RMSE, root mean squared error; VFA, visceral fat area; PBF, percent body fat; TBW, total body water.

Biological age acceleration across cardiometabolic phenotypes. We applied the final Super Learner biological age model to the cardiometabolic validation cohort to estimate calibrated biological age acceleration relative to the healthy reference group. Age acceleration varied substantially across phenotypes and was most pronounced among participants with overweight or obesity (**Figure 4; Supplementary Figure S1; Supplementary Table S7**). The highest mean acceleration was observed in the overweight/obese hypertensive–hyperglycemic group, followed by the overweight/obese normotensive–hyperglycemic and overweight/obese hypertensive–normoglycemic groups. Participants with overweight/obesity who were normotensive and normoglycemic also showed positive acceleration, although the magnitude was lower.

**Figure 4.**
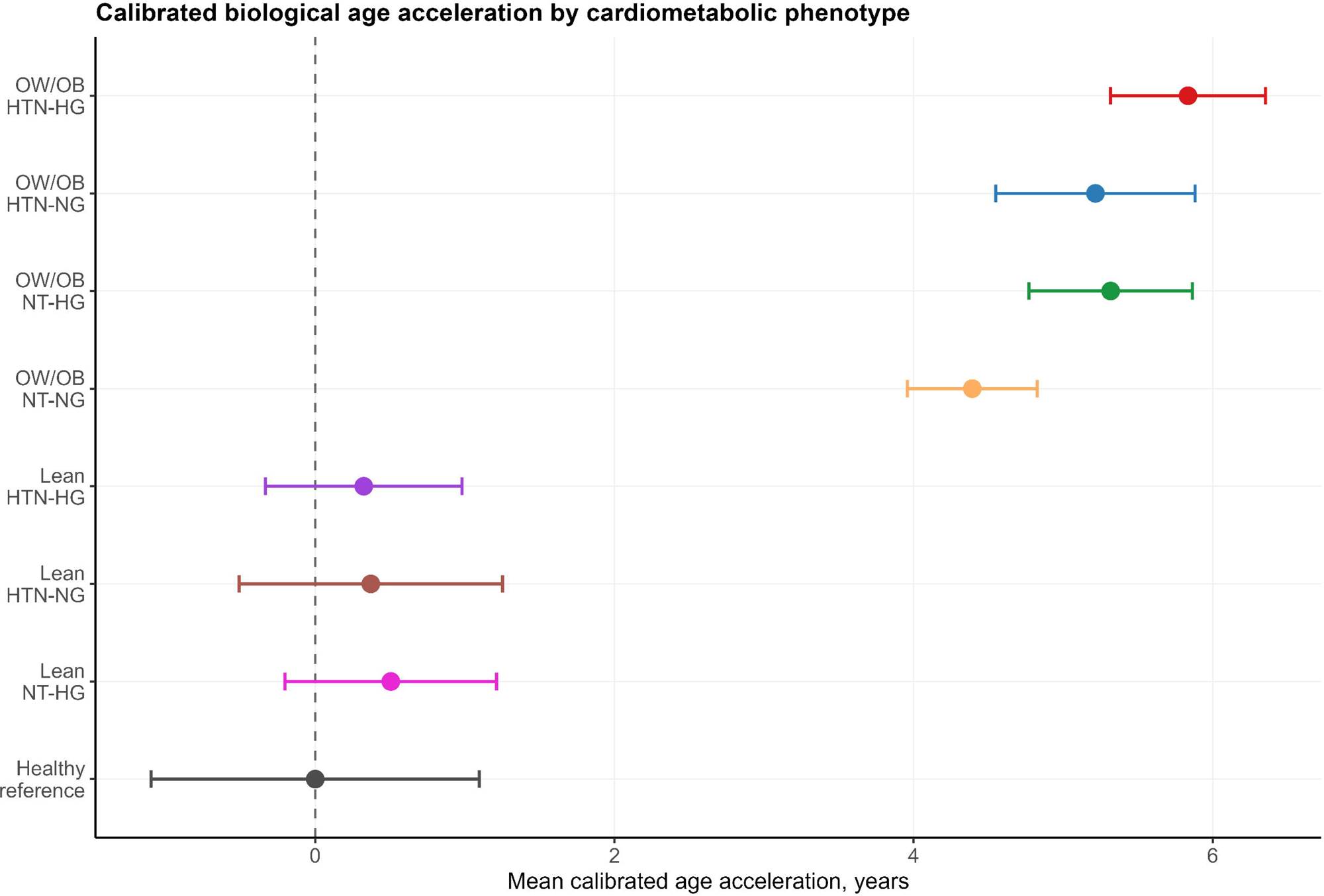
Calibrated biological age acceleration across cardiometabolic phenotypes. Mean calibrated biological age acceleration is shown across cardiometabolic phenotype groups defined by BMI status, blood pressure status, and glycemic status. The lean normotensive-normoglycemic group was used as the healthy reference group. Points indicate group-wise mean calibrated age acceleration, and horizontal error bars indicate 95% confidence intervals. The vertical dashed line at zero represents the expected biological age pattern of the healthy reference group. Positive values indicate higher predicted biological age than expected for chronological age. Abbreviations: OW/OB, overweight/obese; HTN, hypertensive; NT, normotensive; HG, hyperglycemic; NG, normoglycemic.

Mean calibrated age acceleration was 5.83 years in the overweight/obese hypertensive– hyperglycemic group, 5.32 years in the overweight/obese normotensive–hyperglycemic group, 5.22 years in the overweight/obese hypertensive–normoglycemic group, and 4.39 years in the overweight/obese normotensive–normoglycemic group (**Figure 4; Supplementary Table S7**). In contrast, lean cardiometabolic phenotype groups showed mean acceleration values close to the healthy reference: 0.32 years in the lean hypertensive–hyperglycemic group, 0.37 years in the lean hypertensive–normoglycemic group, and 0.50 years in the lean normotensive– hyperglycemic group. Collectively, these findings indicate that the biological age-acceleration signal captured by the final model was concentrated primarily in overweight/obese cardiometabolic phenotypes.

## Discussion

In this study, we developed an inflammation-associated biological age model using circulating cytokines, sex, body-composition measures, and hemoglobin, and applied it across cardiometabolic phenotype groups in a South Asian cohort. The model captured a reproducible, moderate age-related biological signal in the healthy reference cohort and identified marked biological age acceleration among participants with overweight or obesity, particularly when hypertension and hyperglycemia co-occurred. These findings suggest that immune-metabolic biomarker patterns may capture meaningful heterogeneity in biological aging across cardiometabolic risk states[9, 11, 13, 30].

The exploratory cytokine analyses showed that CTACK, MIG, HGF, MIP1a, and MCSF were among the cytokines most positively associated with chronological age. These associations were modest but consistent with the concept that aging involves coordinated low-grade immune and inflammatory changes rather than large shifts in a single cytokine[9, 11]. Notably, each of these cytokines has been implicated independently in immunometabolic aging in other populations. CC-chemokines related to CTACK have been causally linked to ageing-associated traits in Mendelian randomization analyses[31]. MIG contributes to inflammaging scores that predict long-term mortality in older adults[32]. HGF has been shown to be among the plasma proteins most strongly associated with metabolic age across large population cohorts[33]. MIP-1a is upregulated in aging visceral adipose tissue, and M-CSF signaling in monocytes and macrophages is also associated with inflammaging [34, 35].

The sex-stratified concordance analysis further suggested that several cytokine-age relationships were shared between females and males, although some cytokines showed sex-specific variation in effect size. This is consistent with previous demonstrations that the trajectory of inflammaging, despite being shared between the two sexes, may vary in magnitude and timing[36]. While several age-associated cytokines identified in this study reflect components of senescence-associated inflammatory signaling, these biomarkers should be interpreted as correlates of immune aging rather than evidence of causal senescence pathways.

By training the model in lean normotensive-normoglycemic participants, we attempted to learn a reference pattern of immune-metabolic aging under comparatively healthy steady-state physiology. Application of this reference model to cardiometabolic phenotypes allowed us to quantify deviation from this healthy aging pattern. The Super Learner model achieved moderate age-prediction performance in the healthy reference cohort, which is expected given that circulating cytokines and body-composition markers represent only part of the broader biology of aging[16, 31]. The ensemble approach allowed both linear and nonlinear predictor structures to contribute to prediction, with the largest weights assigned to XGBoost and ordinary least-squares regression.

Predictor-contribution analysis indicated that the final model reflected an immune-metabolic signal rather than a cytokine-only inflammatory signal. Visceral fat area and percent body fat were the strongest contributors, followed by cytokines including CTACK and SDF1a, as well as hemoglobin and sex. This finding is biologically plausible because adiposity, particularly visceral adiposity, is closely linked to chronic low-grade inflammation and metabolic dysfunction [37]. Visceral adipose tissue in particular becomes an increasingly important driver of systemic inflammation with age, secreting elevated levels of chemokines that recruit circulating monocytes and amplify local and systemic inflammatory signaling[30]. Therefore, the model is best interpreted as an inflammation-associated or immune-metabolic biological age model, rather than a purely cytokine-derived inflammatory clock.

The strongest biological age acceleration was observed in overweight/obese participants, with the greatest acceleration among those who were both hypertensive and hyperglycemic. Overweight/obese participants who were normotensive and normoglycemic also showed positive acceleration, although of lower magnitude. This suggests that excess adiposity alone may be associated with an older immune-metabolic profile, while additional cardiometabolic abnormalities may further accentuate this signal. In contrast, lean participants with hypertension and/or hyperglycemia showed mean acceleration values close to the healthy reference group, indicating that the model-derived acceleration was driven more strongly by adiposity-related biology than by blood pressure or glycemic status alone.

These findings may be relevant for biological age modeling in South Asian populations, where cardiometabolic risk often emerges at lower BMI thresholds and may be accompanied by distinct body-composition patterns[7, 8]. The present analysis represents an initial step toward developing a South Asian biological age model based on relatively low-cost and scalable immune-metabolic features. Future work should test whether a further reduced panel of cytokines and low-cost clinical or body-composition variables can provide robust biological age estimation across diverse South Asian populations.

The cross-sectional nature of the study poses a major limitation, as biological age acceleration cannot be interpreted as a longitudinal rate of aging. Moreover, the healthy reference cohort was modest in size for high-dimensional biological age modeling. Although we used repeated cross-validation and conservative learner tuning to reduce overfitting, external validation in an independent cohort will be important. Cytokine measurements included exact zero values, which were interpreted as below the assay detection or reporting limit and replaced with a small constant; this preprocessing choice may influence cytokine distributions. Finally, permutation importance reflects predictive contribution, not causal biological importance, particularly because correlated cytokines and body-composition variables may share predictive information.

Overall, this study shows that circulating cytokines and body-composition features capture a measurable immune-metabolic aging signal in a South Asian cohort. The resulting biological age model identified marked acceleration among overweight/obese cardiometabolic phenotypes, especially when hypertension and hyperglycemia co-occurred. These findings support the use of integrated inflammatory and metabolic biomarkers to study heterogeneity in biological aging across cardiometabolic risk states.

## Author Contributions

DG and PT conceptualised the study. PT did the formal data analysis. DG, MG and NB contributed to the data analyses. SP, IS, RR, AV, SRK, SP, VSK, MAU, NR, AS, YK, PHL, AM, VY, APS, AaM, SR, KBT, GRC, MJK, MD, RW, JK, PM, US, SM, PC, KC, SS, VS, SC were responsible for data acquisition, data management and investigations presented within the study. VS, DG and SC were responsible for project administration and resource allocation for data acquisition for the present study. PT, MG and DG wrote the original draft. PT, MG, VS, SC and DG had final responsibility for the decision to submit for publication. All authors approved the final draft of the manuscript.

## Data Availability

All data produced in the present study are available upon reasonable request to the authors.

## Acknowledgement

We acknowledge CSIR-IGIB as the nodal lab of Phenome India Cohort study for logistics and administrative support. Support from assisting zonal labs, directors/heads of all CSIR labs and centers and all coordinators is acknowledged. The monitoring committee is acknowledged for its timely suggestions and course corrections. We acknowledge participants and volunteers. We further acknowledge Phenome India Consortium Study Group (tabulated in Supplementary Appendix 1) and the personnel listed in Supplementary Appendix 2.

---

**Supplementary Appendix 1. Acknowledgement Names**

**Supplementary Appendix 2. Phenome India Consortium Study Group**

**Supplementary Figure S1. Predicted biological age versus observed chronological age across cardiometabolic phenotype groups.**

Faceted scatter plots show predicted biological age from the final Super Learner model versus observed chronological age across cardiometabolic phenotype groups. Points represent individual participants and are distinguished by sex, with solid points indicating females and open points indicating males. Colored lines indicate phenotype-specific linear trends, and dashed grey lines indicate the identity line. Group-specific R² and sample size are shown within each panel. The healthy reference group comprised lean normotensive-normoglycemic participants. Abbreviations: OW/OB, overweight/obese; HTN, hypertensive; NT, normotensive; HG, hyperglycemic; NG, normoglycemic.

**Supplementary Figure S2. Cytokine-age associations across all retained cytokines.**

Multi-page scatter plots show log2 cytokine abundance versus chronological age for each of the 44 retained cytokines. Each panel uses cytokine-specific y-axis scaling to show within-cytokine variation. Points represent individual participants and trend lines are shown by sex. Spearman rho and false discovery rate-adjusted P values are displayed within each panel.

**Supplementary Table S1. Repeated cross-validation performance of candidate age-prediction models.**

Repeated 5-fold cross-validation performance is shown for XGBoost, elastic-net regression, and the Super Learner model, including R², mean absolute error, and root mean squared error.

**Supplementary Table S2. Average Super Learner ensemble weights.**

Mean ensemble weights are shown for each candidate learner included in the Super Learner library.

**Supplementary Table S3. Overall cytokine-age associations.**

Spearman correlations between chronological age and each retained cytokine are shown for the full analytic cohort, with Benjamini-Hochberg false discovery rate correction.

**Supplementary Table S4. Sex-stratified cytokine-age associations.**

Spearman correlations between chronological age and each retained cytokine are shown separately in females and males.

**Supplementary Table S5. Age-and sex-adjusted cytokine differences across cardiometabolic phenotype groups.**

Linear model estimates compare each cardiometabolic phenotype group with the lean normotensive-normoglycemic healthy reference group for each cytokine.

**Supplementary Table S6. Permutation variable importance for the Super Learner biological age model.**

Model-agnostic permutation importance estimates are shown for each predictor, including changes in root mean squared error, mean absolute error, and R² after predictor permutation.

**Supplementary Table S7. Calibrated biological age acceleration by cardiometabolic phenotype group.**

Group-wise calibrated biological age acceleration summaries are shown for each phenotype group, including means, confidence intervals, medians, and interquartile ranges.

