## Supplementary Appendix 1 for "A Scalable Biological Clock for Metabolic Disease Prediction from the Phenome India Cohort"

**Supplementary Appendix 1: Phenome India Consortium Study Group Members (Apart from the listed authors)**

| Sl. No | Names | Surname |
|---|---|---|
| 1 | Abhishek | Kumar |
| 2 | Meghana | Arvind |
| 3 | Ankur | Halder |
| 4 | Anshul | Bhardwaj |
| 5 | Anshuman | Sahu |
| 6 | Ayushi | Narayan |
| 7 | Debasis | Dash |
| 8 | Deeksha | Yadav |
| 9 | Deepak | |
| 10 | Kalyani | Verma |
| 11 | Komal | Jindal |
| 12 | Jafar | Sarif |
| 13 | Mohit | |
| 14 | Pankaj | Pandey |
| 15 | Pranjal | Tewari |
| 16 | Ranit | D'Rozario |
| 17 | Praveena | Mishra |
| 18 | Rohit | Kumar |
| 19 | Purbita | Bandopadhyay |
| 20 | Safeer | Khan |
| 21 | Shail | Kumari |
| 22 | Shilpa | Ray |
| 24 | Shubham | Kumar |
| 25 | Rituparna | Jana |
| 26 | Sumant | Kumar |
| 27 | Swarnendu | Bag |
| 28 | Dipayan | Sarkar |
| 29 | Tanmay | Pawaskar |
| 30 | Tarini | Mathur |
| 31 | Vivek | Junghare |
| 32 | Dipamoy | Dutta |
| 33 | Jahangir | Alam |
| 34 | Pratitusti | Basu |
| 35 | Saheli | Chowdhury |
| 36 | Saikat | Majumder |
| 37 | Dibya Rana Saha | Roy |
| 38 | Jukanti | Akshitha |
| 39 | MK | Kanakavalli |
| 40 | Rakhesh | K V |
| 41 | Ajit A | Sutar |
| 42 | Ameya A | Pawar |

| | | |
|---|---|---|
| 43 | Ankita | Namdeo |
| 44 | Apurva | Balge |
| 45 | Ashok P | Giri |
| 46 | Chiranjit | Chowdhury |
| 47 | Dhanasekaran | Shanmugam |
| 48 | Milind | Kale |
| 49 | Narendra Y | Kadoo |
| 50 | Nikhilesh | Yadav |
| 51 | Rashdajabeen Q | Shaikh |
| 52 | Sagar | Baulia |
| 53 | Shivani V | Palkar |
| 54 | Shrutika M | Shewale |
| 55 | Shyam K | Gawari |
| 56 | Syed G | Dastager |
| 57 | Vaishnavi N | Mahajan |
| 58 | Bhabani S | Jena |
| 59 | Boopathy | Ramasamy |
| 60 | Sai Adarsh | Sahu |
| 61 | Sk Rameej | Raja |
| 62 | T Pavan | Kumar |
| 63 | Trupti | Das |
| 64 | Jagadeshwar Reddy | Thota |
| 65 | Prabhakar | Sripadi |
| 66 | Ramakrishna | Sistla |
| 67 | Ramesh | Ummanni |
| 68 | Sai Balaji | Andugulapati |
| 69 | Srinivasa Rao | M |
| 70 | Adrija | Rakshit |
| 71 | Amit Kumar | Shahravat |
| 72 | Amit | Lahiri |
| 73 | Deepanshu | Sindhwani |
| 74 | Kabita | Sarkar |
| 75 | Kajal | KM |
| 76 | Lakra | Promila |
| 77 | Mrigank | Srivastava |
| 78 | Rahul | Roy |
| 79 | Shail | Singh |
| 80 | Shikha | Yadav |
| 81 | Smita | Pandey |
| 82 | Vivek | Bhosale |
| 83 | Gopal Krishna | Patra |
| 84 | Iranna | Gogeri |
| 85 | Narendra | Singh |
| 86 | Raju | Khan |
| 87 | Neeraj | Jain |
| 88 | Rajesh Kumar | Verma |

| | | |
|---|---|---|
| 89 | Ganesh | Venkatachalam |
| 90 | Murugan | Veerapandian |
| 91 | Amit | Kumar |
| 92 | Deepak | Bansal |
| 93 | Dheeraj Kumar | Kharbanda |
| 94 | Dinesh | Gupta |
| 95 | Sk. Masiul | Islam |
| 96 | Vipul | Sharma |
| 97 | Prakash M | Halami |
| 98 | S P | Muthukumar |
| 99 | Anil Kumar | Maurya |
| 100 | Anirban | Pal |
| 101 | Daneshvar | Prasad |
| 102 | A K | Raman |
| 103 | Bhanu | Pandey |
| 104 | Dikchha | Singh |
| 105 | Jai Krishna | Pandey |
| 106 | Parimala | Karupannan |
| 107 | Suresh Kumar | Anandasadagopan |
| 108 | Vandhana | Anumaiya |
| 109 | Swati | Saha |
| 110 | Vishal | Anand |
| 111 | Mukti | Advani |
| 112 | Rina | Singh |
| 113 | Anamika | Kothari |
| 114 | Suman | Singh |
| 115 | Avinash | Mishra |
| 116 | Pooja | Aggarwal |
| 117 | Shreedhar | Kanagarjan |
| 118 | Ankita | Kumari |
| 119 | Ravi | Raj |
| 120 | Vikram | Patial |
| 121 | Yogendra | Padwad |
| 122 | Fayaz | Malik |
| 123 | Kaneez | Fatima |
| 124 | Nancy | Sharma |
| 125 | Sahaurti | Sharma |
| 126 | Sakshi | Nagial |
| 127 | Sumit G | Gandhi |
| 128 | Debashish | Ghosh |
| 129 | Jyoti | Porwal |
| 130 | Pramod | Chauhan |
| 131 | Suchismita | Benjwal |
| 132 | Neha | Mehrotra |
| 133 | Prabhanshu | Tripathi |
| 134 | Vikas | Srivastava |

| | | |
|---|---|---|
| 135 | Amit | Tuli |
| 136 | Anshu | Bhardwaj |
| 137 | Bhupender | Singh |
| 138 | Deepak | Sharma |
| 139 | Kuldeep | Singh |
| 140 | Lalit | Kumar |
| 141 | Parvez | Ahmad |
| 142 | Pradip | Sen |
| 143 | Pranavathiyani | G |
| 144 | Pravin | Kumar |
| 145 | Priyadarshan | Kinatukara |
| 146 | Priyanshu Singh | Raikwar |
| 147 | Rakesh | Kumar |
| 148 | Rashmi | Kumar |
| 149 | Ritu | Jatav |
| 150 | Shiva Sundharam | S |
| 151 | Siddhakam | Palmal |
| 152 | Simran | Gambhir |
| 153 | Srinivasan | Krishnamurthi |
| 154 | Abbani | Rakesh |
| 155 | Prakash | L |
| 156 | Satisha | Shri |
| 157 | Indrani | Ghosh |
| 158 | Brahma Nanda | Singh |
| 159 | Chandana Venkateswara | Rao |
| 160 | Madan Mohan | Pandey |
| 161 | Sanjeev Kumar | Ojha |
| 162 | Vijayanandraj | Selvaraj |
| 163 | Prashanti | Niwant |
| 164 | Shilpa | Paranjape |
| 165 | Manuj Kr | Das |
| 166 | Pankaj | Bharali |
| 167 | Sukanya | Borkakoti |
| 168 | Tridip | Phukan |
| 169 | Biswajit | Mandal |
| 170 | EVSSK | Babu |
| 171 | T Vijaya | Kumar |
| 172 | Rajeev K | Sukumaran |
| 173 | Rameshkumar | N |
| 174 | Bhumika | Shirodkar |
| 175 | Kalpana Sandesh | Chodankar |
| 176 | Samir Ravikant | Damare |
| 177 | Akshika | |
| 178 | Arun | Uniyal |
| 179 | Arvind | Meena |
| 180 | Ansu J | Kailath |

| | | |
|---|---|---|
| 181 | K Sudhakara | Rao |
| 182 | Krishna | Kumar |
| 183 | Kuldeep Singh | Gour |
| 184 | Navneet Singh | Randhawa |
| 185 | Nikhil | Kumar |
| 186 | Priyanka | Singh |
| 187 | Roshan | Kumar |
| 188 | Arun Kant | Singh |
| 189 | Ved Varun | Agrawal |
| 190 | Maheswaran | Srinivasan |
| 191 | Vasudevan | Pandurangan |
| 192 | Manisha | Sakpal |
| 193 | Rashmi | Arya |
