## Supplementary Appendix 2 for "A Scalable Biological Clock for Metabolic Disease Prediction from the Phenome India Cohort"

| Supplementary Appendix 2 | | | |
|---|---|---|---|
| Sl. No | First Name | Last Name | Institute |
| 1 | Arpana | Parihar | CSIR-AMPRI |
| 2 | Shalu | Yadav | CSIR-AMPRI |
| 3 | Mohd Abubakar | Sadique | CSIR-AMPRI |
| 4 | Akshay Singh | Tomar | CSIR-AMPRI |
| 5 | Varsha | Agrawal | CSIR-AMPRI |
| 6 | Sakshi | Rajput | CSIR-AMPRI |
| 7 | Aabha | Kushwah | CSIR-AMPRI |
| 8 | Shilpee | Chauhan | CSIR-AMPRI |
| 9 | Varsha | Parmar | CSIR-AMPRI |
| 10 | Pushpesh | Ranjan | CSIR-AMPRI |
| 11 | Neeraj | Kumar | CSIR-AMPRI |
| 12 | Abhisek | Giri | CSIR-AMPRI |
| 13 | Navnidhi | Tripathi | CSIR-AMPRI |
| 14 | Jyoti | Lodhi | CSIR-AMPRI |
| 15 | Parul | Shrivastava | CSIR-AMPRI |
| 16 | Prachi | Shrivastava | CSIR-AMPRI |
| 17 | Komal | Sharma | CSIR-AMPRI |
| 18 | Surya Narayan | Mishra | CSIR-CCMB |
| 19 | Sathya | Swetha | CSIR-CCMB |
| 20 | Gururaj | Kalshetti | CSIR-CCMB |
| 21 | Rakesh Pal | Bhagat | CSIR-CCMB |
| 22 | Chandreshwar Raju | Kataru | CSIR-CCMB |
| 23 | Hari Charan Goud | Nerella | CSIR-CCMB |
| 24 | Amit | Chakraborty | CSIR-CCMB |
| 25 | Prangya Paramita | Sahoo | CSIR-CCMB |
| 26 | Alagu | Sankareswaran | CSIR-CCMB |
| 27 | Sofia | Banu | CSIR-CCMB |
| 28 | Priyadarshini | Sanyal | CSIR-CCMB |
| 29 | Sourav | Ganguly | CSIR-CCMB |
| 30 | Pallavi Rao | T | CSIR-CCMB |
| 31 | Akila | Ramesh | CSIR-CCMB |
| 32 | Abhishek | Saha | CSIR-CCMB |
| 33 | Shreya | | CSIR-CCMB |
| 34 | Pramoda | | CSIR-CCMB |
| 35 | Ruqaiya | Tasneem | CSIR-CCMB |
| 36 | Sangavi | Pakkyam | CSIR-CECRI |
| 37 | Khushbu Singh | Raghav | CSIR-CEERI |
| 38 | Shivam | Tiwari | CSIR-CEERI |
| 39 | Vipul | Sharma | CSIR-CEERI |
| 40 | Vishavjeet | | CSIR-CEERI |
| 41 | Garishma | Kalra | CSIR-CEERI |
| 42 | Vikas | Rao | CSIR-CEERI |
| 43 | Jagmohan | Pathak | CSIR-CEERI |
| 44 | Krishana | Saini | CSIR-CEERI |

| | | | |
|---|---|---|---|
| 45 | Deepak Kumar | Panwar | CSIR-CEERI |
| 46 | Yetendra | | CSIR-CEERI |
| 47 | Bibin | Samuel | CSIR-CEERI |
| 48 | Jatin | Yadav | CSIR-CEERI |
| 49 | Akriti | Bajpai | CSIR-CEERI |
| 50 | Sher | Singh | CSIR-CEERI |
| 51 | Biddika | Prudvi | CSIR-CEERI |
| 52 | N | Vinodkumar | CSIR-CFTRI |
| 53 | Avilash Sumithra Rani | C | CSIR-CFTRI |
| 54 | Pramila | Epparti | CSIR-CFTRI |
| 55 | M | Archana | CSIR-CFTRI |
| 56 | Debabrata | Chanda | CSIR-CIMAP |
| 57 | Dnyaneshwar Umrao | Bawankule | CSIR-CIMAP |
| 58 | Pankhuri | Singh | CSIR-CIMAP |
| 59 | Pankaj Kumar | Shukla | CSIR-CIMAP |
| 60 | Parmanand | Kumar | CSIR-CIMAP |
| 61 | Chandra | Kant | CSIR-CIMAP |
| 62 | Sarita | Upadhyay | CSIR-CIMAP |
| 63 | Sumit | Kushwaha | CSIR-CIMAP |
| 64 | Kavita | Singh | CSIR-CIMAP |
| 65 | Srinivetha | Pathmanapan | CSIR-CLRI |
| 66 | Syed Nasar | Rahaman | CSIR-CLRI |
| 67 | Vikash | Negi | CSIR-CLRI |
| 68 | Jegan | K | CSIR-CLRI |
| 69 | Sankar | Palanivel | CSIR-CLRI |
| 70 | Lishadevi | M | CSIR-CLRI |
| 71 | Pranathy | K | CSIR-CLRI |
| 72 | Kirubakaran | BJ | CSIR-CLRI |
| 73 | Thiyaneshwar | | CSIR-CLRI |
| 74 | Sujata | Pachhal | CSIR-CMERI |
| 75 | Sukanta | Maji | CSIR-CMERI |
| 76 | Sonali | Garai | CSIR-CMERI |
| 77 | Darshan | Singh | CSIR-CRRI |
| 78 | Mukesh | Kumar | CSIR-CRRI |
| 79 | Sakshi | Gupta | CSIR-CRRI |
| 80 | Keshav | Kaushik | CSIR-CRRI |
| 81 | Satyajit | Nayak | CSIR-CRRI |
| 82 | Lakshmy | T | CSIR-CRRI |
| 83 | Sandeep | Kumar | CSIR-CSIO |
| 84 | Rajni | | CSIR-CSIO |
| 85 | Preetismita | Borah | CSIR-CSIO |
| 86 | Manish | Kumar | CSIR-CSIO |
| 87 | Goraj | Singh | CSIR-CSIO |
| 88 | Suraj | Prakash | CSIR-CSIO |
| 89 | Ajay | Kumar | CSIR-CSIO |
| 90 | Inderjit | | CSIR-CSIO |

| | | | |
|---|---|---|---|
| 91 | Shavita | | CSIR-CSIO |
| 92 | Jyoti | Jangra | CSIR-CSIO |
| 93 | Aman | Grewal | CSIR-CSIO |
| 94 | Shivalika | | CSIR-CSIO |
| 95 | Rajrani | | CSIR-CSIO |
| 96 | Munish | | CSIR-CSIO |
| 97 | Prashant | Kumar | CSIR-CSIO |
| 98 | Satinder | Kaur | CSIR-CSIO |
| 99 | Parneet | Kaur | CSIR-CSIO |
| 100 | Abhishek | | CSIR-CSIO |
| 101 | Salwa | Naushin | CSIR-IGIB |
| 102 | Subhash | Gurjar | CSIR-IGIB |
| 103 | Rajesh | Kumar | CSIR-IGIB |
| 104 | Anushka | | CSIR-IGIB |
| 105 | Aditya | Ranout | CSIR-IHBT |
| 106 | Trilok | Saini | CSIR-IHBT |
| 107 | Sahdev | Choudhary | CSIR-IHBT |
| 108 | Amit | Kumar | CSIR-IHBT |
| 109 | Shweta | Sharma | CSIR-IHBT |
| 110 | Neha | Bhardwaj | CSIR-IHBT |
| 111 | Kajal | Kalia | CSIR-IHBT |
| 112 | Suresh | Kumar | CSIR-IHBT |
| 113 | Abhishek | Goel | CSIR-IHBT |
| 114 | Ravi | Kumar | CSIR-IHBT |
| 115 | Surbhi | Mali | CSIR-IHBT |
| 116 | Komal | Goel | CSIR-IHBT |
| 117 | Swati | Katoch | CSIR-IHBT |
| 118 | Shagun | Dogra | CSIR-IHBT |
| 119 | Vinesh | Sharma | CSIR-IHBT |
| 120 | Rajneesh | Kumar | CSIR-IHBT |
| 121 | Poonam | Dhiman | CSIR-IHBT |
| 122 | Pardeep | Poonia | CSIR-IHBT |
| 123 | Sahiba | Chahal | CSIR-IHBT |
| 124 | Rahul | Kumar | CSIR-IHBT |
| 125 | Athrinandan S. | Hegde | CSIR-IHBT |
| 126 | Rupinder | Kaur | CSIR-IHBT |
| 127 | Vivek | Kumar | CSIR-IHBT |
| 128 | Prakriti | Sharma | CSIR-IHBT |
| 129 | Mohak | Mali | CSIR-IHBT |
| 130 | Priyanka | Koundal | CSIR-IHBT |
| 131 | Sheetal | | CSIR-IHBT |
| 132 | Nidhi | Sharma | CSIR-IICT |
| 133 | Anjali | Veeram | CSIR-IICT |
| 134 | Komal | walvekar | CSIR-IICT |
| 135 | Suriya | panneer | CSIR-IICT |
| 136 | Hari Priya | Sripadi | CSIR-IICT |

| | | | |
|---|---|---|---|
| 137 | Vaishnavi | Kambhampati | CSIR-IICT |
| 138 | Harleen | Kaur | CSIR-IIP |
| 139 | Ayan | Banerjee | CSIR-IIP |
| 140 | Rahul | Gautam | CSIR-IIP |
| 141 | Mirnalini | Juyal | CSIR-IIP |
| 142 | Mridul | Budakoti | CSIR-IIP |
| 143 | Preetanshika | Tracy | CSIR-IIP |
| 144 | Pratima Ashoke | Patel | CSIR-IIP |
| 145 | Ritu | Mourya | CSIR-IIP |
| 146 | Harish | Panwar | CSIR-IIP |
| 147 | Pramod | Chauhan | CSIR-IIP |
| 148 | Sneha | Mohanty | CSIR-IITR |
| 149 | Mohd Tauseef | Khantwal | CSIR-IITR |
| 150 | Vaibhavi | Lahane | CSIR-IITR |
| 151 | Shreya | Tripathi | CSIR-IITR |
| 152 | Sunita | Devi | CSIR-IITR |
| 153 | Gulafsha | Siddiqui | CSIR-IITR |
| 154 | Km | Priya | CSIR-IITR |
| 155 | Priyanka | Goswami | CSIR-IITR |
| 156 | Sandip | Chatterjee | CSIR-IITR |
| 157 | Sakshi | Singh | CSIR-IITR |
| 158 | Sachin | Mishra | CSIR-IITR |
| 159 | Devendri | Khantwal | CSIR-IITR |
| 160 | Ankita | Das | CSIR-IMMT |
| 161 | Mayuri | Kumari | CSIR-IMMT |
| 162 | Amit Kumar | Swain | CSIR-IMMT |
| 163 | Nilotpal | Kapri | CSIR-IMMT |
| 164 | Ghrutanjali | Sahu | CSIR-IMMT |
| 165 | Suchismita | Senapati | CSIR-IMMT |
| 166 | Subhashree | Nayak | CSIR-IMMT |
| 167 | Yogesh | Chaudhary | CSIR-IMMT |
| 168 | Anagonou | Bertrand | CSIR-IMMT |
| 169 | Bibekananda | Nayak | CSIR-IMMT |
| 170 | Saswati | Suryasnata | CSIR-IMMT |
| 171 | Annapurna | Behera | CSIR-IMMT |
| 172 | Satyabrata | Sahoo | CSIR-IMMT |
| 173 | Akshyurna | Pattnaik | CSIR-IMMT |
| 174 | Rakhi Rani | Biswas | CSIR-IMMT |
| 175 | Chandrama | Dipa | CSIR-IMMT |
| 176 | Adyasha Bijay | Mishra | CSIR-IMMT |
| 177 | Dinesh Kumar | Panda | CSIR-IMMT |
| 178 | Sudipta | Nayak | CSIR-IMMT |
| 179 | Avishek | Kar | CSIR-IMMT |
| 180 | Kajal | Sundaray | CSIR-IMMT |
| 181 | Swagatika | Dash | CSIR-IMMT |
| 182 | Alok | Pattanaik | CSIR-IMMT |

| | | | |
|---|---|---|---|
| 183 | Sonali Swapanjali | Bhoi | CSIR-IMMT |
| 184 | Swikruti | Mishra | CSIR-IMMT |
| 185 | Shalini | Das | CSIR-IMMT |
| 186 | Parinita | Mishra | CSIR-IMMT |
| 187 | Debidatta | Barik | CSIR-IMMT |
| 188 | Tejaswini | Das | CSIR-IMMT |
| 189 | Subhashree | Pattnaik | CSIR-IMMT |
| 190 | Kailash T | Bhamare | CSIR-IMTECH |
| 191 | Chander Shekhar | Sharma | CSIR-IMTECH |
| 192 | Amit | Kumar[1] | CSIR-IMTECH |
| 193 | Vineet | Kumar | CSIR-IMTECH |
| 194 | Deepak | Bhatt | CSIR-IMTECH |
| 195 | Surjeet | Singh | CSIR-IMTECH |
| 196 | Harminder | Singh | CSIR-IMTECH |
| 197 | Paramjit | Lal | CSIR-IMTECH |
| 198 | Sandeep | Kumar | CSIR-IMTECH |
| 199 | Nitin | Sharma | CSIR-IMTECH |
| 200 | Jaideep | Mehta | CSIR-IMTECH |
| 201 | Renu | | CSIR-IMTECH |
| 202 | RK | Dhiman | CSIR-IMTECH |
| 203 | Anjali | Koundal | CSIR-IMTECH |
| 204 | Bhumika | Vaidya | CSIR-IMTECH |
| 205 | Amit | Kumar[2] | CSIR-IMTECH |
| 206 | Shweta | Pandey | CSIR-IMTECH |
| 207 | Anunay | Sinha | CSIR-IMTECH |
| 208 | Shubham | | CSIR-IMTECH |
| 209 | Bulbul | Roy | CSIR-IMTECH |
| 210 | Pompi | Bhadra | CSIR-IMTECH |
| 211 | Meena | Sharma | CSIR-IMTECH |
| 212 | Sana | Khatun | CSIR-IMTECH |
| 213 | Payal | Thakur | CSIR-IMTECH |
| 214 | Bhagyashree | Rabha | CSIR-IMTECH |
| 215 | Meenal | Rastogi | CSIR-IMTECH |
| 216 | Mayur Sudhakar | Zarkar | CSIR-IMTECH |
| 217 | Somnath | Chindhe | CSIR-IMTECH |
| 218 | Shreya | Singh | CSIR-IMTECH |
| 219 | Neelam | | CSIR-IMTECH |
| 220 | Rakesh | Kumar | CSIR-IMTECH |
| 221 | Siddhakam | Palmal | CSIR-IMTECH |
| 222 | Lalit | Kumar | CSIR-IMTECH |
| 223 | Joyasree | Das | CSIR-IMTECH |
| 224 | Parvez | Ahmad | CSIR-IMTECH |
| 225 | Kiran Raj | S | CSIR-IPU |
| 226 | Ravi Kumar | Sharma | CSIR-IPU |
| 227 | Nataraj | D | CSIR-NAL |
| 228 | Swetha C | Desai | CSIR-NAL |

| | | | |
|---|---|---|---|
| 229 | Shivanna | TB | CSIR-NAL |
| 230 | Murthy | BYK | CSIR-NAL |
| 231 | Shashidhara | KN | CSIR-NAL |
| 232 | Raghavendra Swamy | M | CSIR-NAL |
| 233 | Aravind Kumar | Joseph | CSIR-NAL |
| 234 | Purushotham | HM | CSIR-NAL |
| 235 | Muniprakash | M | CSIR-NAL |
| 236 | Shekhar | G | CSIR-NAL |
| 237 | Ranganatha | S | CSIR-NAL |
| 238 | SatishKumar | Shri | CSIR-NAL |
| 239 | Shashikala | U | CSIR-NAL |
| 240 | Praveen Kumar | JD | CSIR-NAL |
| 241 | Nikhilesh | Yadav | CSIR-NCL |
| 242 | Parveen | Goyal | CSIR-NCL |
| 243 | NMR | Ashwin | CSIR-NCL |
| 244 | Shashikala | Ranjane | CSIR-NCL |
| 245 | Priyanka M | Bankar | CSIR-NCL |
| 246 | Amreen | Sheikh | CSIR-NCL |
| 247 | Shahaji | Palaskar | CSIR-NCL |
| 248 | Jugal | Kanerkar | CSIR-NCL |
| 249 | Arvind | Chourasiya | CSIR-NCL |
| 250 | Bhagyashree | Likhitkar | CSIR-NCL |
| 251 | Pradeep | S | CSIR-NCL |
| 252 | Pranay | Awathare | CSIR-NCL |
| 253 | Swaraj | Jathar | CSIR-NCL |
| 254 | Yugendra | Patil | CSIR-NCL |
| 255 | Rohit S | Dashpute | CSIR-NCL |
| 256 | Minal R | Bhalerao | CSIR-NCL |
| 257 | Vinod | Kamble | CSIR-NCL |
| 258 | Yashodhara | Shinde | CSIR-NCL |
| 259 | Vineetkumar S | Nair | CSIR-NCL |
| 260 | Prathmesh | Ghongade | CSIR-NCL |
| 261 | Prashant | Kalaskar | CSIR-NCL |
| 262 | Ganesh | Jadhav | CSIR-NCL |
| 263 | Preshitta | Bhat | CSIR-NCL |
| 264 | Tanaji B | Devkate | CSIR-NCL |
| 265 | Vikram | Nichit | CSIR-NCL |
| 266 | Sonali | Gaikwad | CSIR-NCL |
| 267 | Anand Kumar | Shukla | CSIR-NCL |
| 268 | Sakshi R | Mangate | CSIR-NCL |
| 269 | Ameya A | Pawar | CSIR-NCL |
| 270 | Ankita | Namdeo | CSIR-NCL |
| 271 | Disha | Dwivedi | CSIR-NCL |
| 272 | Apurva | Barge | CSIR-NCL |
| 273 | Pankaj | Pardhi | CSIR-NEERI |
| 274 | Jayashree Chiring | Phukon | CSIR-NEIST |

| | | | |
|---|---|---|---|
| 275 | Shridhar | Hiremath | CSIR-NEIST |
| 276 | Prachurjya | Dutta | CSIR-NEIST |
| 277 | Parishmita | Borgohain | CSIR-NEIST |
| 278 | Moirangthem Goutam | Singh | CSIR-NEIST |
| 279 | Devpratim | Koch | CSIR-NEIST |
| 280 | Dipanneeta Das | Gupta | CSIR-NEIST |
| 281 | Pankaj | Barman | CSIR-NEIST |
| 282 | Sahana | SK | CSIR-NEIST |
| 283 | Monojit Kumar | Roy | CSIR-NEIST |
| 284 | Aditya | Sarkar | CSIR-NEIST |
| 285 | Bhaben | Sharma | CSIR-NEIST |
| 286 | Shyamalima | Mech | CSIR-NEIST |
| 287 | Masoom | Saikia | CSIR-NEIST |
| 288 | Trishna Rani | Borah | CSIR-NEIST |
| 289 | Gayatri | Gogoi | CSIR-NEIST |
| 290 | Anupriya | Borah | CSIR-NEIST |
| 291 | Udeshna | Changmai | CSIR-NEIST |
| 292 | Himadri | Das | CSIR-NEIST |
| 293 | Anshuman | Goswami | CSIR-NEIST |
| 294 | Rocktotpal | Mahanta | CSIR-NEIST |
| 295 | Rina | Yumnam | CSIR-NEIST |
| 296 | Sukanya | Borthakur | CSIR-NEIST |
| 297 | Manabendra | Borah | CSIR-NEIST |
| 298 | Sarangthem Dinamani | Singh | CSIR-NEIST |
| 299 | Pronami | Gogoi | CSIR-NEIST |
| 300 | Priyanka | Saikia | CSIR-NEIST |
| 301 | Umakanta | Tanti | CSIR-NEIST |
| 302 | Mohan | Kurmi | CSIR-NEIST |
| 303 | Ravi Kumar | Sahu | CSIR-NEIST |
| 304 | Nayan Jyoti | Borah | CSIR-NEIST |
| 305 | Babli | Borah | CSIR-NEIST |
| 306 | Sarifa Mafuz | Ahmed | CSIR-NEIST |
| 307 | Bidyut Prakash | Deka | CSIR-NEIST |
| 308 | Darshana | Tamuli | CSIR-NEIST |
| 309 | Meghna | Kakoty | CSIR-NEIST |
| 310 | Bhaswati V | Borah | CSIR-NEIST |
| 311 | Yashodhara | Goswami | CSIR-NEIST |
| 312 | Priyanka | Boro | CSIR-NEIST |
| 313 | Inshamol | KP | CSIR-NIIST |
| 314 | Athulya | | CSIR-NIIST |
| 315 | Amrutha | M | CSIR-NIIST |
| 316 | Anaswara P | A | CSIR-NIIST |
| 317 | Anupama | | CSIR-NIIST |
| 318 | Aparna | | CSIR-NIIST |
| 319 | Aparna | Dinil | CSIR-NIIST |
| 320 | Archa R | S | CSIR-NIIST |

| | | | |
|---|---|---|---|
| 321 | Archana | | CSIR-NIIST |
| 322 | Arnold | Moses | CSIR-NIIST |
| 323 | Athira V | C | CSIR-NIIST |
| 324 | Biji | Raphy | CSIR-NIIST |
| 325 | Dileep R | Nair | CSIR-NIIST |
| 326 | Greeshma | | CSIR-NIIST |
| 327 | Karthika | Nath | CSIR-NIIST |
| 328 | Lakshmi M | Nair | CSIR-NIIST |
| 329 | Lakshmi | Shaji | CSIR-NIIST |
| 330 | Neetha P | Sobandas | CSIR-NIIST |
| 331 | Reena | R | CSIR-NIIST |
| 332 | Saranyadevi | | CSIR-NIIST |
| 333 | Shehbas | C | CSIR-NIIST |
| 334 | Valan | Rebinro | CSIR-NIIST |
| 335 | Adarsh | VP | CSIR-NIIST |
| 336 | Govind | Ranade | CSIR-NIO |
| 337 | AS | Unnikrishnan | CSIR-NIO |
| 338 | Sohan Pal | Meena | CSIR-NIO |
| 339 | Karishma Pradeep | Chari | CSIR-NIO |
| 340 | Vitasta | Jad | CSIR-NIO |
| 341 | Vikash | Kumar | CSIR-NIO |
| 342 | Nishamol | M | CSIR-NIO |
| 343 | Helen | Agnes | CSIR-NIO |
| 344 | Natasha Maria | Barnes | CSIR-NIO |
| 345 | Bhumika Rohidas | Shirodkar | CSIR-NIO |
| 346 | Shania Waluscha | Moeres | CSIR-NIO |
| 347 | Yuvrani | Halarnkar | CSIR-NIO |
| 348 | Ramila Ram | Gaonkar | CSIR-NIO |
| 349 | Mrinalini Chandra | Mohan | CSIR-NIO |
| 350 | Pratibha | Bachhley | CSIR-NIO |
| 351 | Himani | Meena | CSIR-NIScPR |
| 352 | Abhinav | Banait | CSIR-NIScPR |
| 353 | Shekar | Sharma | CSIR-NIScPR |
| 354 | Rajesh | | CSIR-NPL |
| 355 | Sumana | Gajjala | CSIR-NPL |
| 356 | Sudesh | Yadav | CSIR-NPL |
| 357 | Vinod Kumar | Tanwar | CSIR-NPL |
| 358 | Vishesh | Garg | CSIR-NPL |
| 359 | Manoj Kumar | Pandey | CSIR-NPL |
| 360 | Vikash | Sharma | CSIR-NPL |
