## Supplementary figures and images for "A Scalable Biological Clock for Metabolic Disease Prediction from the Phenome India Cohort"

### Supplementary Figure 1

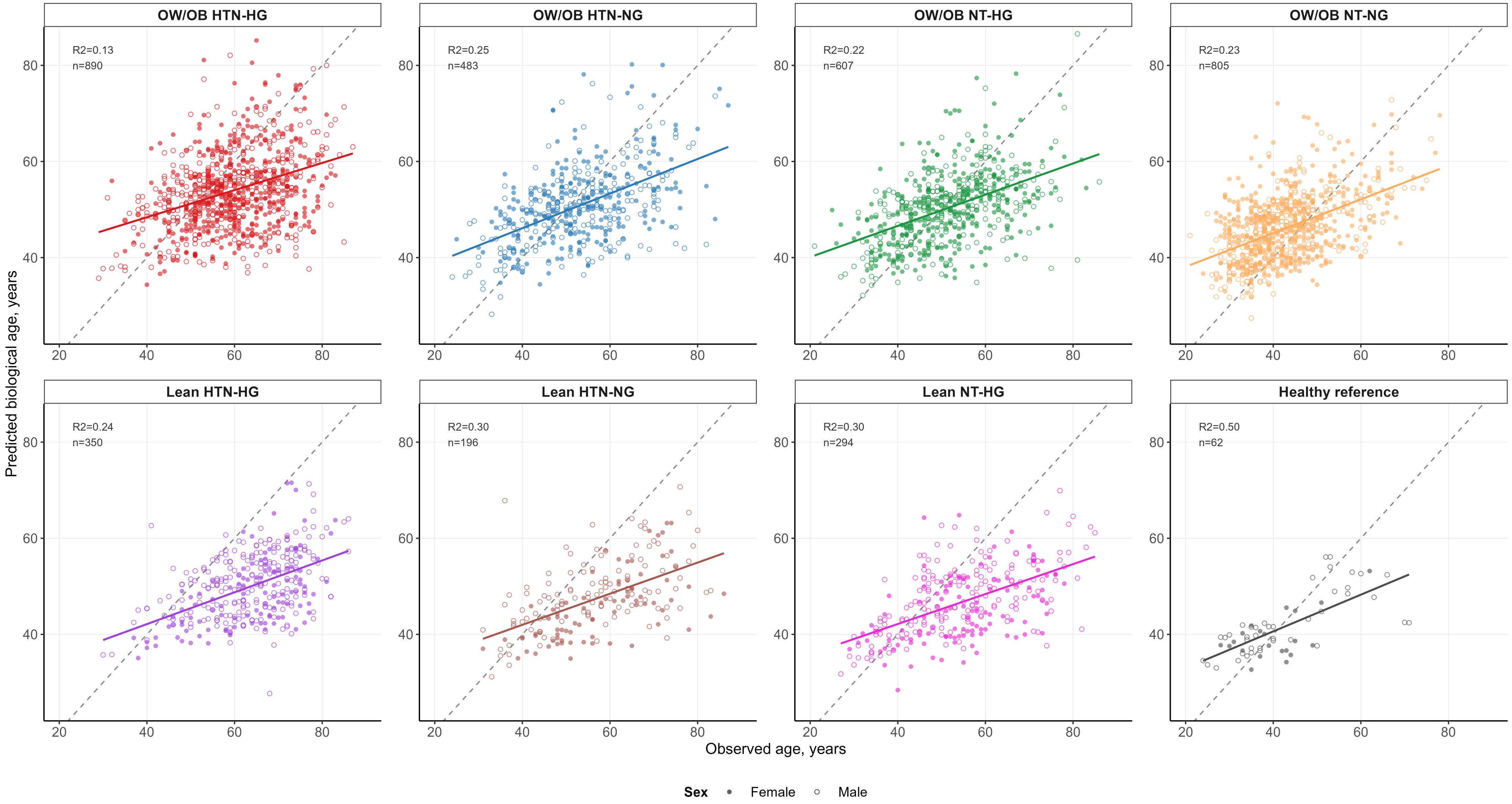

### Supplementary Figure 2

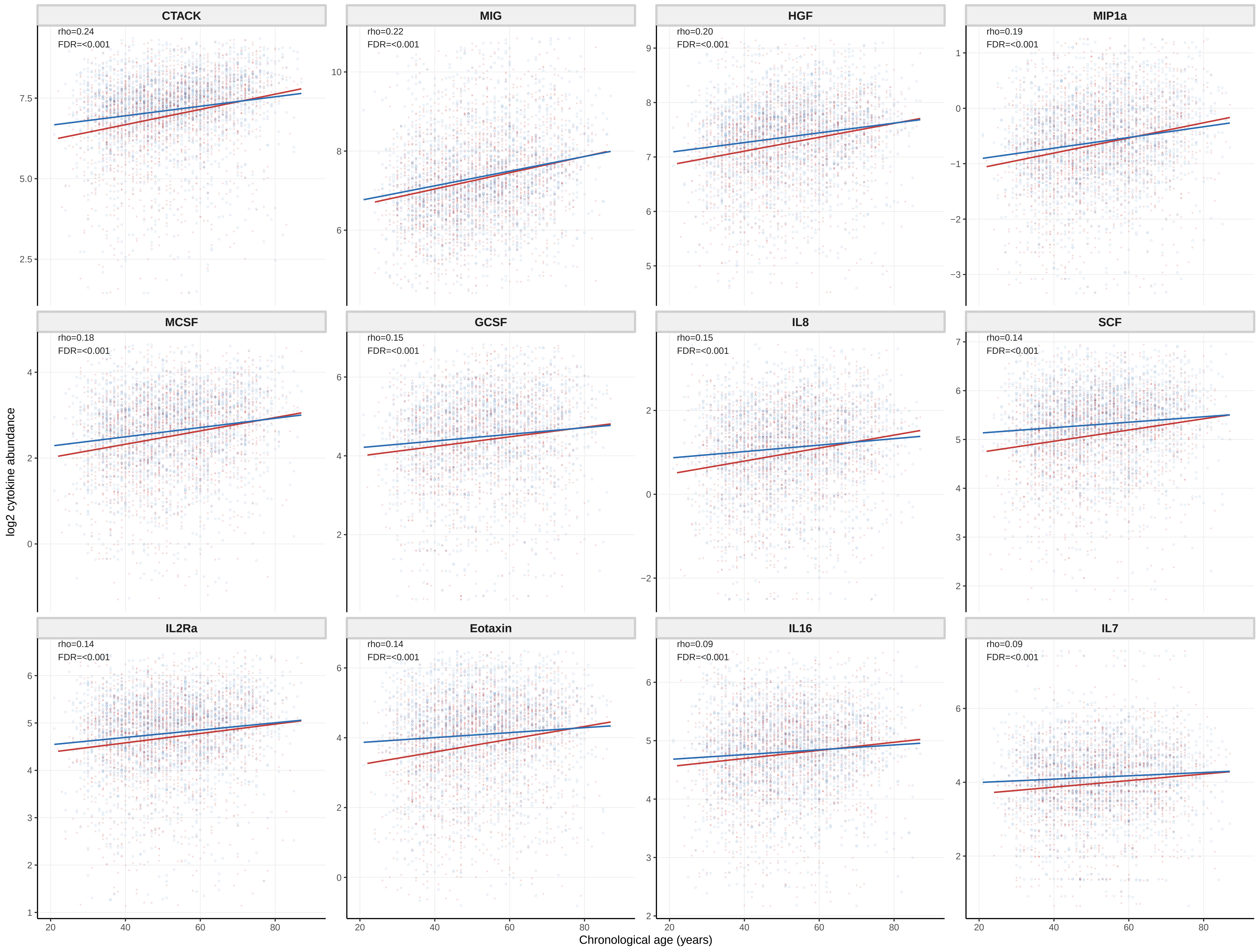

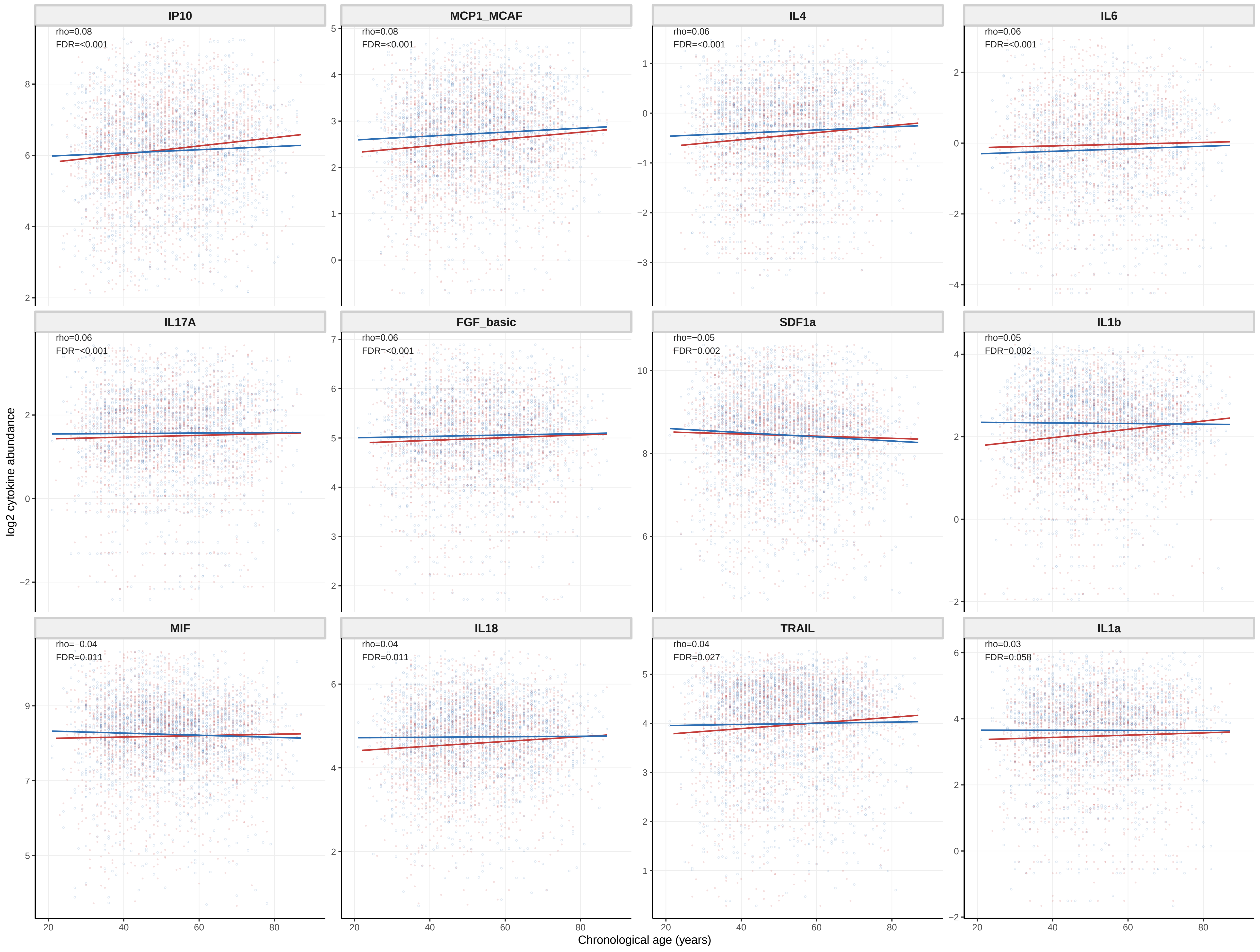

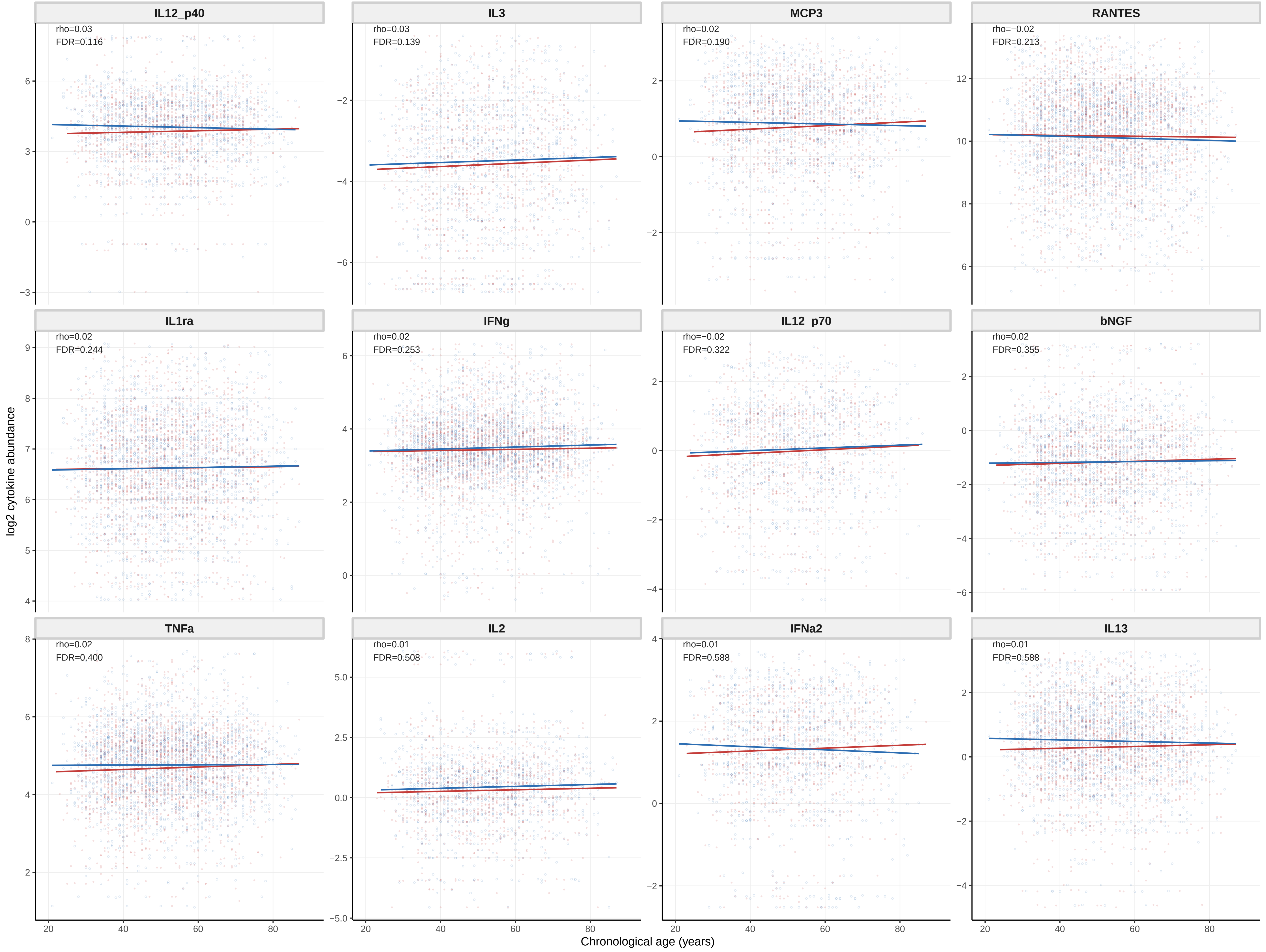

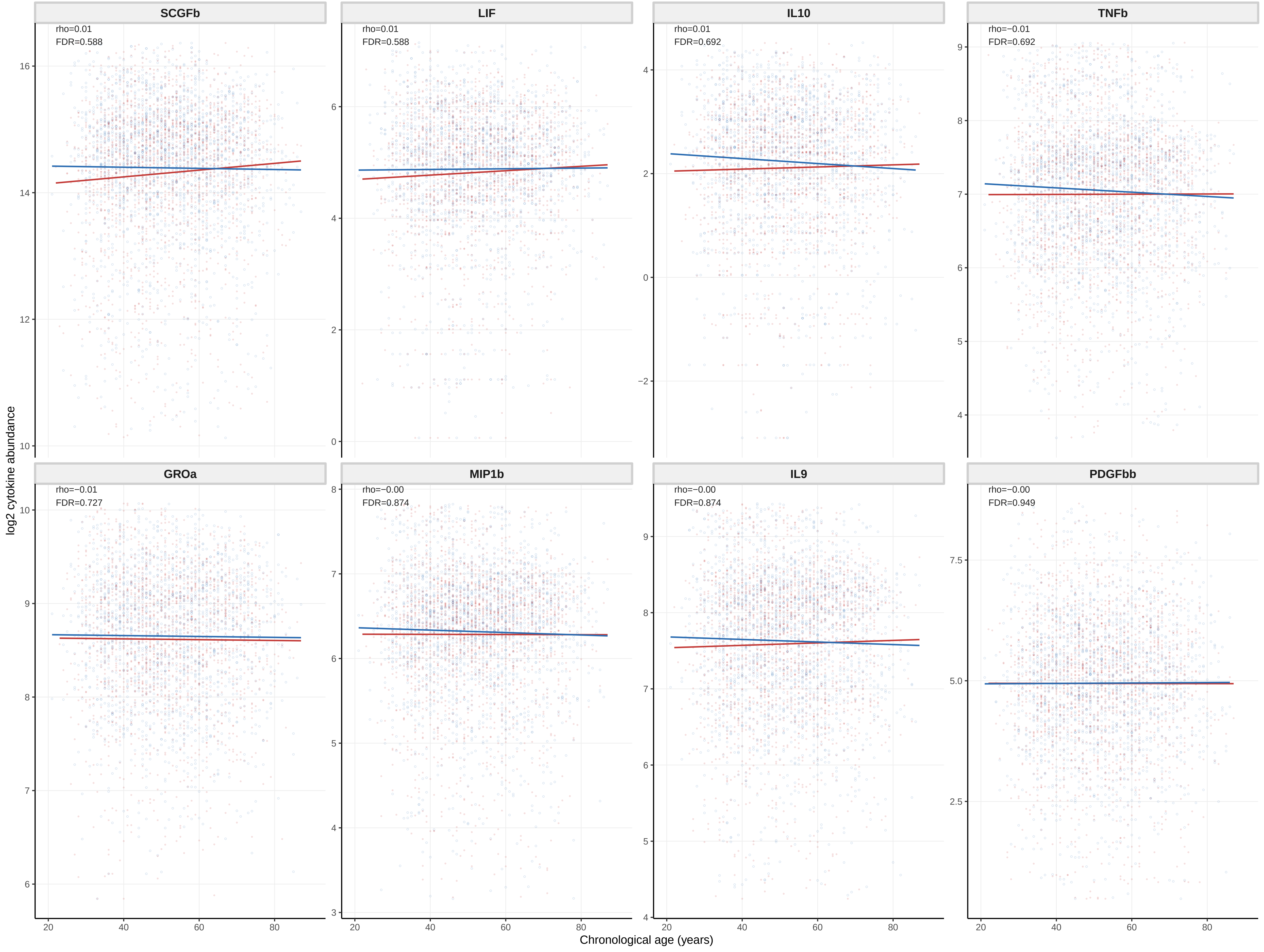
